# Cardiovascular risk and hippocampal-cognitive coupling in Alzheimer’s disease

**DOI:** 10.64898/2026.06.01.26354601

**Authors:** Sofia Fernandez-Lozano, Sylvia Villeneuve, D. Louis Collins, the Alzheimer’s Disease Neuroimaging Initiative

**Affiliations:** Montreal Neurological Institute, McConnell Brain Imaging Centre, Montreal, Quebec, Canada; Department of Neurology and Neurosurgery, McGill University, Montreal, Quebec, Canada; Department of Psychiatry, McGill University, Montreal, Quebec, Canada; StoP-AD Centre, Douglas Mental Health Institute Research Centre, Montreal, Quebec, Canada; Department of Biomedical Engineering, McGill University, Montreal, Quebec, Canada

**Keywords:** Cardiovascular risk, Framingham Risk Score, hippocampus, cognitive decline, Alzheimer’s disease, MIMIC model, age-confounding

## Abstract

**INTRODUCTION:** The Framingham Risk Score (FRS) indexes cardiovascular risk (CVR), but age-weighting may confound associations with brain and cognitive outcomes.

**METHODS:** In 923 amyloid-positive ADNI participants, we compared FRS against a Multiple Indicators Multiple Causes (MIMIC)-derived age-adjusted measure (CVR_mimic_) using sex-stratified linear mixed-effects (LME) and latent growth curve mediation (LGCM) models of hippocampal-to-ventricle ratio (HVR)– cognitive coupling.

**RESULTS:** FRS predicted hippocampal atrophy in all six LGCM models; CVR_mimic_ in none of the six. HVR–cognitive coupling held in four of six FRS and four of six CVR_mimic_ models. Indirect effects reached significance in four of six FRS and none of the six CVR_mimic_ models. LME 3-way interactions (years × risk × HVR) survived FDR correction in all six FRS versus none of the six CVR_mimic_ models.

**DISCUSSION:** FRS “effects” on hippocampal-cognitive decline largely reflect age-related variance. Age-adjusted measures complement FRS by isolating cardiovascular effects from aging.

**Research in Context:** *Systematic Review:* The Framingham Risk Score (FRS) predicts brain atrophy and cognitive decline across cohorts [1–4]. Yet age dominates the FRS [5,6] and accounts for most of its predictive value in older adults [7,8], suggesting these associations may reflect age confounding. No prior study has compared FRS against an age-adjusted latent measure.

*Interpretation:* FRS indirect effects on cognitive decline via hippocampal atrophy primarily reflect age-weighting; age is itself a legitimate vascular proxy [9]. Partialling out age (CVR_mimic_) nullified the cardiovascular-risk-to-hippocampal pathway, while HVR-cognitive coupling persisted, indicating coupling reflects neurodegeneration. Prior FRS-brain reports likely conflate age-driven and cardiovascular effects.

*Future Directions:* Replication in independent aging cohorts with longitudinal cardiovascular measurement is needed. The MIMIC age-adjustment framework can be applied to any composite risk score where age confounding is a concern. Intervention trials should test whether cardiovascular management preserves hippocampal-cognitive coupling using age-adjusted endpoints rather than FRS.

**Highlights:**

- Age-adjusted cardiovascular risk does not accelerate hippocampal atrophy
- Hippocampal-cognitive coupling persists regardless of how risk is measured
- FRS-brain associations in elderly samples primarily reflect age-weighting
- A reverse pattern in amyloid-negative controls supports a cognitive reserve effect
- MIMIC-based age-adjustment generalizes to any age-confounded composite score

## 1 Background

Cardiovascular risk (CVR) factors are increasingly recognized as modifiable contributors to cognitive decline and dementia risk [10–15]. The Framingham Risk Score (FRS), a widely used composite measure of 10-year risk of cardiovascular event [5] — originally designed and validated for midlife application — has been associated with reduced brain volumes, compromised white matter integrity, and accelerated cognitive decline across multiple aging cohorts [1–3,16]. However, the FRS is heavily weighted by age, with log-hazard coefficients of 3.06 for men and 2.33 for women [5], making age the dominant predictor in the composite, with different effects by sex. In older samples where age variance is restricted, this creates substantial confounding between FRS and any age-related outcome [6,17].

Age is itself a strong proxy for cumulative vascular exposure [5,9], and the FRS age-weighting therefore carries legitimate clinical information. However, this creates an interpretive challenge when FRS is used to predict age-related outcomes: because age simultaneously drives FRS through its formula coefficients and independently drives neurodegenerative outcomes through biological aging, it acts as a common cause of both the predictor and the outcome. Simply controlling for age in regression models does not fully resolve this confound, because age is not an external confounder but an internal component of the score itself. In elderly cohorts such as ADNI, this problem is compounded by the fact that the ADSP-PHC-derived FRS variable is capped at 30% 10-year risk [5,18], compressing variance in the upper range and further limiting sensitivity.

To address this, we employed a Multiple Indicators Multiple Causes (MIMIC) model [19,20], which treats age as a *cause* of cardiovascular risk rather than an indicator. The structural equation CVR = *γ* · Age_*z*_ + ζ (where CVR denotes the latent cardiovascular risk factor) partitions observed cardiovascular risk into an age-explained component (*γ* · Age_*z*_) and a residual (ζ) that captures cardiovascular risk independent of age [21]. The resulting factor scores, termed CVR_mimic_, are orthogonal to age by construction (r ≈ 0), enabling a direct test of whether FRS-brain associations reflect true cardiovascular pathophysiology or age-related confounding.

The primary objective of this study was to determine whether FRS effects on hippocampal-cognitive coupling persist when age-confounding is removed. We tested three hypotheses. First, if FRS effects on hippocampal atrophy are driven by age-weighting, CVR_mimic_ should show a null or attenuated effect of cardiovascular risk on hippocampal atrophy rate (the CVR_mimic_ → hippocampal-to-ventricle ratio [HVR] slope path) in latent growth curve mediation models (LGCM). Second, if FRS mediation is age-confounded, CVR_mimic_ indirect effects through HVR decline should likewise be null. Third, even when mediation is absent, CVR_mimic_ may still moderate the brain-cognition relationship via a three-way interaction (years from baseline [YRS] × CVR [either FRS or CVR_mimic_] × HVR) in linear mixed-effects (LME) models, reflecting a reserve-like mechanism whereby cardiovascular burden modulates the coupling between hippocampal structure and cognitive performance without directly accelerating atrophy. All analyses were stratified by sex, given established sex differences in cardiovascular risk profiles, brain aging trajectories, and AD pathophysiology [22,23].

## 2 Methods

### 2.1 Study Design and Participants

Data used in the preparation of this article were obtained from the Alzheimer’s Disease Neuroimaging Initiative (ADNI) database (adni.loni.usc.edu). The ADNI was launched in 2003 as a public-private partnership, led by Principal Investigator Michael W. Weiner, MD. The primary goal of ADNI has been to test whether serial magnetic resonance imaging (MRI), positron emission tomography (PET), other biological markers, and clinical and neuropsychological assessment can be combined to measure the progression of mild cognitive impairment (MCI) and early Alzheimer’s disease (AD). ADNI data were downloaded from the LONI-IDA portal between December 2025 and February 2026 (latest download: 10 February 2026).

Data were drawn from ADNI phases 1, GO, 2, and 3. We included only amyloid-positive participants, defined by PET Centiloid > 25 [24] across florbetapir (FBP), florbetaben (FBB), and Pittsburgh Compound B (PIB) tracers, or CSF amyloid-positive status via Gaussian Mixture Model classification [25]. A participant was classified as amyloid-positive if positivity was established at any available timepoint (PET or CSF); once classified, all longitudinal visits for that participant were included. Additional inclusion criteria required available MRI-derived hippocampal and ventricular volumes [26], cardiovascular risk factor data (FRS components) [18,27], and at least one cognitive assessment with Alzheimer’s Disease Sequencing Project Phenotype Harmonization Consortium (ADSP-PHC) harmonized composite scores [28]. The resulting sample encompassed the full spectrum of amyloid-positive individuals, including cognitively unimpaired (CU), mild cognitive impairment (MCI), and Alzheimer’s disease dementia (AD). Participant inclusion and exclusion flow is shown in Figure 1.

**Figure 1:**
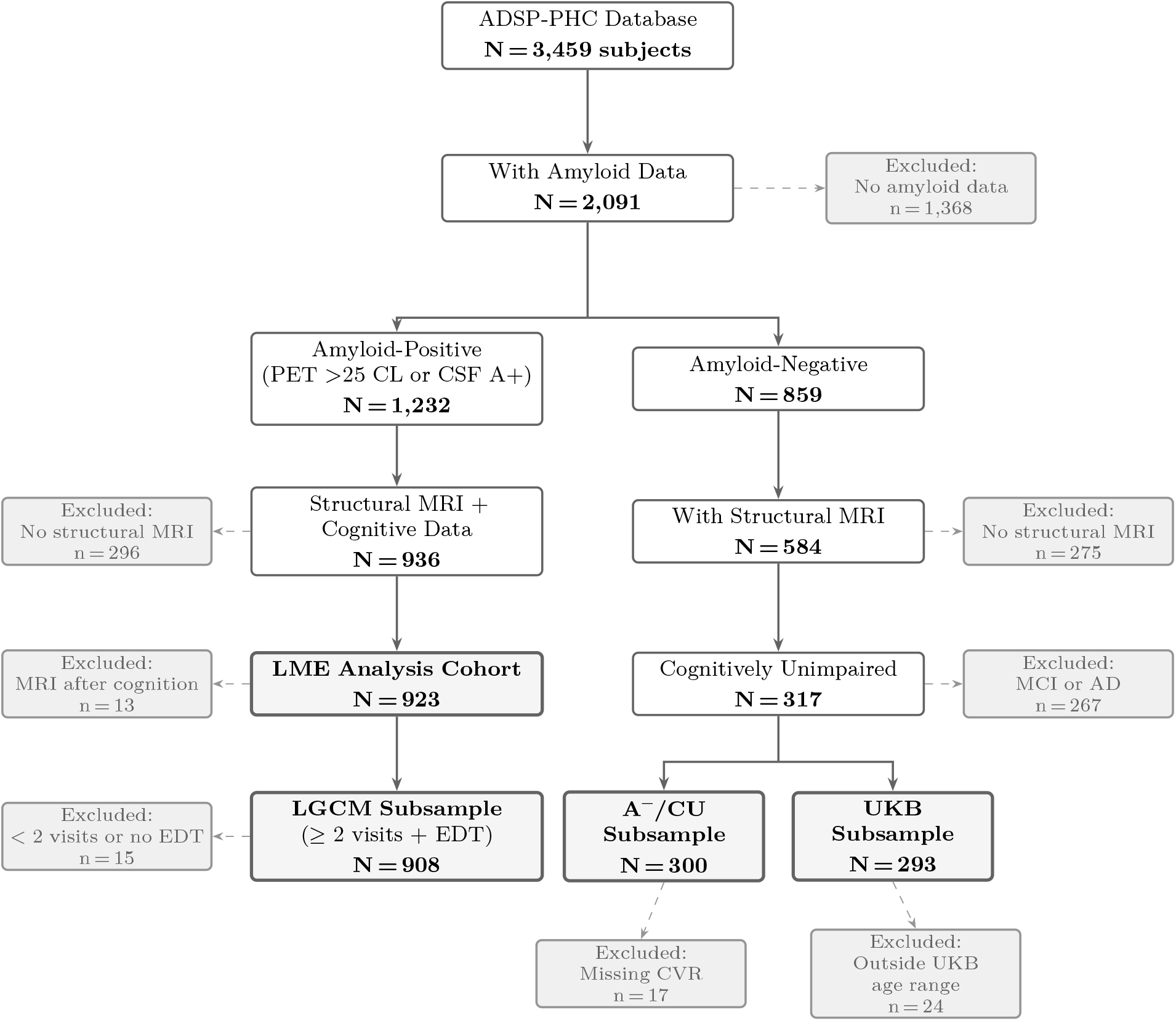
CONSORT flow diagram showing participant selection. Left: amyloid-positive analysis track from LME cohort to LGCM subsample. Right: amyloid-negative, cognitively unimpaired participants split into the A^−^/CU sensitivity subsample and the UKB normative comparison subsample. ADSP-PHC = Alzheimer’s Disease Sequencing Project Phenotype Harmonization Consortium; CL = Centiloid; CSF = cerebrospinal fluid; CU = cognitively unimpaired; EDT = estimated disease time; MRI = magnetic resonance imaging; UKB = UK Biobank.

A separate normative comparison sample of amyloid-negative, cognitively unimpaired ADNI participants with structural MRI within the UK Biobank (UKB) age range (46–82 years) was used to validate the trans-fer of UKB-derived z-scores to the ADNI cohort (see Normative Transfer Validation). Note that UKB participants are population-based without amyloid testing; the normative GAMLSS (Generalized Additive Models for Location, Scale, and Shape) models therefore reflect the general population rather than a confirmed amyloid-negative sample. The UKB normative reference is sex-imbalanced (56.2% female), reflecting the source imaging cohort and quality-control filtering [29]; sex-stratified GAMLSS fitting ensures within-sex z-score calibration is unaffected by this imbalance.

### 2.2 Neuroimaging: Hippocampal-to-Ventricle Ratio (HVR)

T1-weighted magnetization-prepared rapid gradient echo (MPRAGE) sequences were acquired on 1.5T and 3T scanners across ADNI sites using standardized acquisition protocols [30]. Hippocampal (HC) and temporal horn of the lateral ventricle (VC) volumes were obtained using a convolutional neural network (CNN) trained on manual labels [26], which demonstrated low failure rates in ADNI compared to multi-atlas and patch-based alternatives. Volumes were provided in stereotaxic space (mm^3^) and converted to native space via subject-specific scale factors.

We defined the hippocampal-to-ventricle ratio (HVR) as:

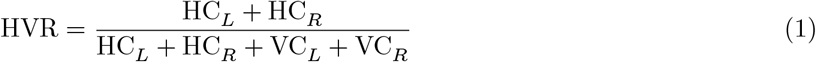

computed on bilateral summed native-space volumes [31]. This ratio captures hippocampal integrity relative to surrounding cerebrospinal fluid (CSF)-filled spaces, where hippocampal atrophy is accompanied by compensatory expansion of the temporal horns (i.e., ex vacuo dilation). HVR approaches 1.0 when hippocampal volume is large relative to the temporal horns (preserved structure) and approaches 0.0 when the hippocampus is small relative to the temporal horns (atrophy with ventricular expansion). The ratio inherently normalizes for head size without requiring intracranial volume (ICV) correction, as both numerator and denominator scale proportionally with head size [29].

HVR z-scores were derived from Generalized Additive Models for Location, Scale, and Shape (GAMLSS; [32]) fit on UKB data (N = 27,434 cognitively healthy adults, age 45–82). The distribution family was Box-Cox Cole and Green (BCCG), which models three distributional parameters — location (*μ*, the conditional median), scale (*σ*, the coefficient of variation), and skewness (*v*, the Box-Cox power) — as smooth functions of covariates: age (penalized cubic spline), education (linear), and intracranial cavity volume (ICC), with separate models fit for each sex. The resulting z-score quantifies how many standard deviations an individual’s observed HVR deviates from the conditional median predicted by the GAMLSS model given their age, sex, education, and head size. Negative z-scores indicate disproportionate hippocampal volume loss relative to the normative reference population. These are cross-sectional deviation scores, not measures of longitudinal change rate.

#### 2.2.1 Normative Transfer Validation

Valid cross-cohort application of UKB-trained GAMLSS models requires verification that z-scores are calibrated in the target ADNI population. We performed validation in a comparable ADNI subsample of amyloid-negative, cognitively unimpaired participants within the UKB age range (46–82 years), where z-scores would be expected to approximate a standard normal distribution (mean zero, standard deviation one). We compared mean z-scores, standard deviations, and 95% confidence intervals against expected *N*(0, 1) and visualized age-bin-specific calibration via forest plot.

### 2.3 Cardiovascular Risk Assessment

#### 2.3.1 Framingham Risk Score (FRS)

We obtained FRS values from the Alzheimer’s Disease Sequencing Project Phenotype Harmonization Consortium (ADSP-PHC) Cardiovascular Risk Factor dataset [18,27]. The ADSP-PHC computed FRS using the Framingham 2008 simple office-based 10-year cardiovascular disease (CVD) risk score via the `ascvd_10y_frs_simple` function from the CVrisk R package [33]. Despite the function name, this implements the Framingham general cardiovascular risk algorithm [5], not the American College of Cardiology/American Heart Association (ACC/AHA) atherosclerotic cardiovascular disease (ASCVD) Pooled Cohort Equations. The score estimates 10-year cardiovascular disease risk based on sex, age (≥ 30 years), body mass index (BMI, kg/m^2^), systolic blood pressure (mmHg), use of blood pressure medication, current smoking status, and diabetes status. This is the “simple” FRS variant that substitutes BMI for the lipid panel (total and high-density lipoprotein [HDL] cholesterol) required by the general FRS [5]. Age coefficients in the log-hazard scale are 3.06 (men) and 2.33 (women), making age the dominant component in older adults. The ADSP-PHC-derived variable is capped at 30% 10-year risk, creating a ceiling effect in elderly samples.

#### 2.3.2 MIMIC Model for Age-Adjusted CVR (CVR_mimic_)

We specified the CVR_mimic_ latent factor as a Multiple Indicators Multiple Causes (MIMIC) model [19,20], in which age is an exogenous cause of the latent cardiovascular risk factor and the disturbance ζ captures residual cardiovascular risk independent of age [21].

##### 2.3.2.1 Model Specification

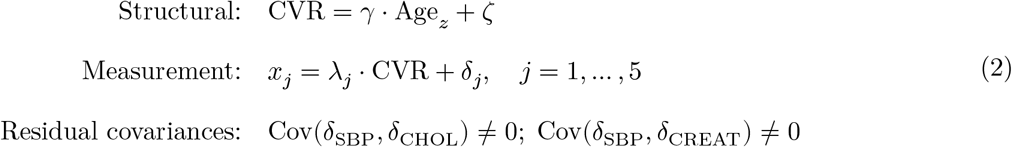

The model includes five measurement indicators spanning vascular, metabolic, and renal cardiovascular risk domains: systolic blood pressure (SBP; z-scored, continuous), hypertension (history of hypertension; binary), fasting glucose (z-scored, continuous), total cholesterol (z-scored, continuous), and serum creatinine (z-scored, continuous). We specified two correlated residuals: SBP ↔ Cholesterol, reflecting shared vascular stiffness and lipid metabolism pathways whereby arterial compliance links blood pressure regulation to lipid profiles beyond the general cardiovascular risk factor; and SBP ↔ Creatinine, reflecting the renal-cardiovascular axis whereby kidney function directly influences blood pressure regulation through the renin-angiotensin system, partially independent of overall cardiovascular risk. Both modification indices and substantive physiological reasoning supported these specifications.

##### 2.3.2.2 Indicator Selection

We determined the final set of five indicators through iterative model refinement, evaluating factor loadings, modification indices, and substantive interpretability at each step. Several candidate indicators were evaluated but excluded. BMI was dropped because its factor loading became non-significant once residual covariances with hypertension, glucose, and cholesterol were modeled, indicating its cardiovascular risk variance is fully captured by the other retained indicators. Diabetes, heart disease, and stroke (ADSP-PHC binary medical history flags) showed near-perfect tetrachoric correlations due to a harmonization artifact in how self-reported conditions were coded across ADNI phases, rendering them empirically indistinguishable. Smoking status had a non-significant factor loading, likely reflecting survivorship bias in an elderly cohort. Blood pressure medication showed near-complete redundancy with hypertension status. Specific loading estimates and model selection diagnostics are reported in the supplement.

##### 2.3.2.3 Estimation and Fit

We evaluated model fit with the robust maximum likelihood (MLR)-scaled comparative fit index (CFI), Tucker-Lewis index (TLI), root mean square error of approximation (RMSEA), and standardized root mean square residual (SRMR).

##### 2.3.2.4 Score Extraction and Age Adjustment

We extracted factor scores in three steps. First, we obtained predicted latent factor scores (*η*) from the fitted MIMIC model via empirical Bayes modal estimation. Second, we computed age-adjusted cardiovascular risk scores (ζ, hereafter CVR_mimic_) as ζ = *η* − *γ* × Age_c_, where *γ* is the estimated regression of the latent cardiovascular risk factor on centered age from the structural path; this removes the age-predicted component of the latent variable, ensuring CVR_mimic_ represents residual cardiovascular risk after partialling out age at the latent level rather than at the indicator level. Third, we z-standardized the resulting ζ scores on the common analysis sample (participants with complete data for both FRS components and MIMIC model indicators) for comparability with FRS. This latent-level residualization operates on the measurement-error-free latent variable and is equivalent to extracting the disturbance term of the MIMIC structural equation [34]. By construction, *r*(CVR_mimic_, Age) ≈ 0.

##### 2.3.2.5 Measurement Invariance

We tested measurement invariance across sex in four steps — configural, metric, scalar, and strict — using the confirmatory factor analysis (CFA) measurement model only (without the Age → cardiovascular risk structural path). Measurement invariance evaluates whether factor loadings, intercepts, and residual variances are equivalent across groups, which is required before interpreting sex-stratified analyses. We evaluated invariance with the Satorra-Bentler scaled chi-square difference test [35], which correctly handles the non-subtractable nature of MLR-scaled test statistics, and with change in fit indices (|ΔCFI| ≤ 0.01, ΔRMSEA ≤ 0.015). When the two criteria disagreed, we adopted the stricter result.

#### 2.3.3 CVR_mimic_ Validation

We validated CVR_mimic_ by examining its correlation with age (expected ≈ 0, confirming successful age-partialling), correlation with FRS (moderate expected, reflecting shared cardiovascular indicators), distribution properties (approximate standard normal), and FRS ceiling analysis (percentage at the 30% risk cap) to document variance compression in the elderly sample.

#### 2.3.4 Cardiovascular Risk Stratification

For trajectory visualizations, we defined CV risk groups by median split of baseline FRS (for FRS analyses) or baseline CVR_mimic_ (for CVR_mimic_ analyses), and similarly defined HVR groups by median split of baseline HVR z-score. Groups were assigned at baseline and held constant across follow-up.

#### 2.3.5 Cardiovascular Risk Predictor Standardization

We z-scored both FRS and CVR_mimic_ on the common analysis sample (defined above) prior to all downstream analyses. This ensures that regression coefficients from both linear mixed-effects (LME) models and latent growth curve models (LGCM) are on an identical scale (effect per 1 SD change), enabling direct comparison of effect magnitudes between FRS and CVR_mimic_. We retained raw (pre-standardized) FRS values for descriptive statistics and ceiling analyses.

### 2.4 Disease Time Estimation

Calendar time (years from baseline) assumes all participants progress at the same rate, which is unrealistic in a heterogeneous AD sample spanning CU, MCI, and dementia stages. Estimated Disease Time (EDT) aligns participants on a common disease timeline by estimating each individual’s position along an underlying progression curve with respect to AD dementia diagnosis [36]. We computed EDT using the progmod R package with a simultaneous multivariate model fitting the Alzheimer’s Disease Assessment Scale-Cognitive subscale (ADAS-Cog) 13 and the Mini-Mental State Examination (MMSE) jointly, sharing a common disease time random effect across scales [37]. The simultaneous approach provides more stable individual time estimates than univariate models because it borrows concordant information across cognitive instruments. EDT values were mean-centered for numerical stability while retaining the years metric; centering does not affect between-subject differences in disease timing. For participants with missing EDT at later timepoints (due to dropout), values were extrapolated using a hybrid approach: a linear slope computed from the individual’s observed EDT values when at least two were available, or the sex-stratified population mean EDT slope otherwise, to minimize attrition bias from listwise deletion.

### 2.5 Cognitive Assessment

We used ADSP-PHC composite scores [28] for three cognitive domains: memory (episodic memory composite: Wechsler Memory Scale-Revised [WMS-R] Logical Memory, Rey Auditory Verbal Learning Test [AVLT], ADAS-Cog word recall, MMSE/Montreal Cognitive Assessment [MoCA] memory items), language (language/semantic composite: category fluency, Boston Naming Test, ADAS-Cog naming/commands), and executive function (executive composite: Trail Making A/B, Wechsler Adult Intelligence Scale-Revised [WAIS-R] Digit Span/Symbol, clock drawing). Scores were co-calibrated across multiple cohorts via a bifactor item banking approach, initially across ADNI, Adult Changes in Thought (ACT), and Religious Orders Study/Memory and Aging Project (ROS/MAP), with subsequent addition of the National Alzheimer’s Co-ordinating Center (NACC). Higher composite scores indicate better cognitive performance across all three domains. Each score has an associated standard error (SE) from the CFA scoring model, reflecting measurement precision at each observation. In the LGCM, cognitive composites enter as single indicators with residual variances fixed to SE^2^ (the squared standard error), embedding known measurement error directly from the ADSP-PHC model rather than estimating it as a free parameter.

### 2.6 Statistical Analysis

#### 2.6.1 Analytical Framework

All primary analyses (LME and LGCM) were run twice: once with FRS and once with CVR_mimic_ as the cardiovascular risk predictor. The key test is whether FRS effects disappear when replaced by age-adjusted CVR_mimic_, which would implicate age-confounding rather than true cardiovascular pathophysiology. The central question is whether cardiovascular risk influences the hippocampal-cognitive relationship, and whether the answer depends on how cardiovascular risk is measured.

All models were sex-stratified (each sex analyzed separately), following the National Institutes of Health policy on Sex as a Biological Variable [38]. Sex differences in AD pathophysiology, cardiovascular risk profiles, and brain aging trajectories motivate stratification rather than inclusion of sex as a covariate [22,23]. Stratification allows effect sizes, variance components, and covariate relationships to differ freely between sexes; sex is omitted from covariates because it defines the strata. Covariates in all models include age at baseline, education (years), and Apolipoprotein E (APOE) *ε*4 carrier status.

Primary inferences are based on uncorrected per-test p-values given the pre-specified, hypothesis-driven nature of the analyses [39,40]. As a supplementary check, we applied FDR correction (Benjamini-Hochberg) within each cardiovascular risk measure across the 6 LME tests (3 domains × 2 sexes). FRS and CVR_mimic_ were corrected separately because they address distinct hypotheses. Whether effects surviving uncorrected thresholds also survive FDR is reported in Results. The key inferential comparison (a-path FRS vs a-path CVR_mimic_) is inherently directional and does not require correction across independent model comparisons [40]. For the LGCM mediation models (12 tests: 2 sexes × 3 domains × 2 predictors), no FDR correction was applied: inference rests on the consistency of the a-path pattern across models rather than on any single test.

##### Analytical roadmap

The two modelling frameworks address distinct questions. The LME tests *moderation*: does cardiovascular risk change the HVR-cognition coupling over time, assessed via the 3-way YRS (years from baseline) × CVR × HVR interaction? The LGCM tests *mediation*: does cardiovascular risk drive cognitive decline through hippocampal atrophy, assessed via the indirect path cardiovascular risk → HVR slope → cognitive slope? Comparing FRS and CVR_mimic_ across both frameworks isolates the role of age-weighting in observed cardiovascular risk-brain associations.

##### Formal predictor comparison

Because FRS and CVR_mimic_ were fitted on the same participants, paired tests are required to compare coefficients. In the LGCM, shared-seed bootstrap ensures identical resamples for both predictors, yielding a paired 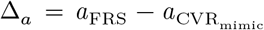 with bias-corrected 95% CI. In the LME, clustered bootstrap (resampling subjects with replacement) provides a paired 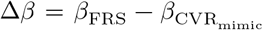 for the 3-way interaction, with bias-corrected 95% CI.

#### 2.6.2 Methodological Validations

##### 2.6.2.1 EDT Linearity Validation

We used likelihood ratio tests (LRT) with LME models comparing linear vs quadratic EDT specifications, testing whether adding a quadratic EDT term improves model fit. This complements the LGCM quadratic convergence findings (see below).

##### 2.6.2.2 Simulation Study: Standard Error-Constrained

Validation

A Monte Carlo simulation validated the SE-constrained LGCM specification (residual variance fixed to SE^2^) against free estimation (residual variance freely estimated). This is a methodological validation, not a power analysis — it validates the estimation approach under known truth, not detection rates for specific effect sizes. The SE-constraint is a key methodological innovation: cognitive composites from the ADSP-PHC CFA have known observation-specific SEs, so fixing residual variance to SE^2^ uses this information rather than discarding it.

Data were generated from a univariate latent growth model with parameters calibrated from the observed ADNI LGCM parallel process models (averaged across all sex × domain combinations): intercept mean = 0 (z-scored composites), slope mean = −0.19 per year EDT (population-average cognitive decline rate), intercept variance = 0.17 (between-person variability in baseline level), slope variance = 0.002 (between-person variability in decline rate), HVR → Slope true effect = 0.12 (mean b-path coupling estimate), measurement SE = 0.29 (median ADSP-PHC composite SE, SD = 0.1 across observations), and EDT with mean = 8.5 years, SD = 5, change = −1 per visit (observed EDT distribution and inter-visit spacing). To assess robustness, the two most consequential parameters — the HVR effect and slope variance — were varied at 0.5×, 1×, and 1.5× their observed values, yielding 9 simulation conditions. Two approaches were compared across 500 replications per condition at N = 200 and N = 500: (a) residual variance fixed to SE^2^ (SE-constrained) and (b) residual variance freely estimated. Performance metrics include parameter bias (mean difference from truth), RMSE (root mean squared error), and 95% CI coverage probability.

#### 2.6.3 LME Growth Models with 3-Way Interaction

The model specification (per sex) is:

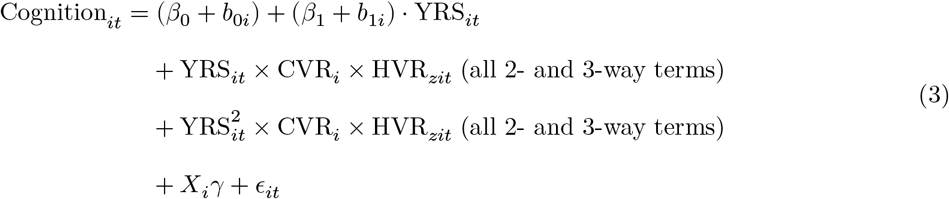

where the linear block (YRS × CVR × HVR) expands to all lower-order terms (YRS × CVR, YRS × HVR, CVR × HVR, and the individual main effects) plus the 3-way interaction; the quadratic block (YRS^2^ × CVR × HVR) similarly expands. R’s * formula operator generates each product term exactly once — two-way terms shared across blocks are not duplicated. The quadratic interaction terms test whether cardiovascular risk moderation of the HVR-cognition relationship accelerates (or decelerates) over follow-up. Random effects include subject-specific intercepts (*b*_0*i*_) and linear slopes (*b*_1*i*_). Cardiovascular risk is entered as either FRS or CVR_mimic_ in parallel model sets. The hypothesis tested is whether cardiovascular risk moderates how the HVR-cognition relationship evolves over time; a negative 3-way *β* means higher cardiovascular risk increasingly weakens the protective HVR-cognition association over follow-up. Inclusion of the quadratic time term (YRS^2^) was evaluated via likelihood ratio tests comparing nested linear and quadratic models across all 12 specifications (2 sexes × 2 brain measures × 3 domains); results are reported in Appendix B (Table B2).

#### 2.6.4 LGCM Parallel Process Models

We specified LGCM with parallel HVR and cognitive trajectories [41,42] to test whether cardiovascular risk affects the hippocampal-cognitive relationship via mediation (CVR → HVR slope → Cognitive slope).

##### 2.6.4.1 Model Specification

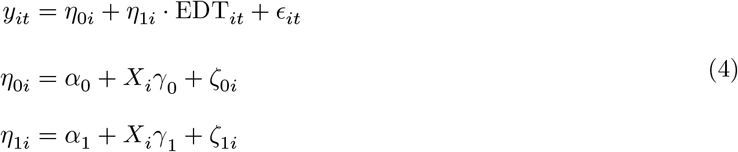

where *y*_*it*_ is the observed score (HVR z-score or cognitive composite) for person *i* at occasion *t*; *η*_0*i*_ and *η*_1*i*_ are person-specific intercepts and slopes; EDT_it_ is the individually-varying time score (see below); *ϵ*_*it*_ is the occasion-specific residual; and *X*_*i*_ includes cardiovascular risk predictor, age, education, and APOE *ε*4 status.

Two parallel processes (HVR and cognitive composite) are specified simultaneously, with their latent growth factors allowed to covary or be linked by directional paths depending on the model, across up to 6 measurement occasions per participant. Cognitive composite residual variances were fixed to SE^2^ from the ADSP-PHC CFA scoring model, embedding known heteroscedastic measurement error directly rather than estimating it as a free parameter; this yields more precise latent slope estimates by separating true variance from measurement noise. For HVR, occasion-specific residual variances were freely estimated; a likelihood ratio test against an equality-constrained alternative (single residual variance pooled across timepoints) rejected the constraint (Δ*χ*^2^ = 35.0, df = 5, *p* < 10^−5^; Male × Memory × CVR_mimic_ cell), consistent with the biologically expected heteroscedasticity of brain atrophy variance over disease time [43].

We first attempted fully random quadratic models (6 latent factors per domain); when these did not converge, we adopted a fixed-quadratic specification: two additional phantom latent factors (HVR quadratic, cognitive quadratic) with variance fixed to near zero and free population means, capturing population-level curvature (2 additional free parameters) without individual-level quadratic variation. Likelihood ratio tests comparing this fixed-quadratic model against a purely linear model determined whether quadratic curvature improved fit. We also attempted to embed the full MIMIC measurement model within the OpenMx parallel process framework; convergence was insufficient, so we used pre-computed ζ scores instead (see Score Extraction above).

##### 2.6.4.2 Time Metric

Slope loadings use individually-varying time scores determined by each participant’s EDT, which maps each visit to a position on the disease progression timeline rather than calendar time. Because individually-varying time scores give each participant a unique implied covariance structure, standard fit indices (CFI, TLI, RMSEA) that require a common saturated reference model cannot be computed [44,45].

##### 2.6.4.3 Covariates and Predictors

The two cardiovascular risk predictors (FRS and CVR_mimic_) were mean-centered separately for males and females to account for sex differences in cardiovascular risk distributions; covariates (age, education) were centered on grand means pooled across sexes for consistency across sex-stratified analyses; APOE *ε*4 carrier status entered as a binary indicator. CVR_mimic_ entered as pre-computed age-adjusted ζ scores from the MIMIC model, z-standardized on the common sample. Both FRS and CVR_mimic_ are observed covariates in the LGCM. We regressed all covariates on both the latent intercept and latent slope factors for each process (HVR and cognitive) and fit the same models with FRS and CVR_mimic_ for direct comparison of predictive utility. Although HVR z-scores are age-normalized via GAMLSS and CVR_mimic_ is age-orthogonal by construction, baseline age was retained as a covariate because cognitive composites are not age-normalized and because normative models trained on one cohort may not fully absorb age effects when applied to a demographically different sample [46,47].

##### 2.6.4.4 Model Fit and Inference

Missing data were handled via full information maximum likelihood (FIML), which uses all available observations without listwise deletion. We assessed model fit via −2 log-likelihood (−2LL), Akaike information criterion (AIC), and Bayesian information criterion (BIC). For bootstrap inference, we used case resampling with n = 1,000 iterations for all mediation paths (a, b, c’, indirect, total), reporting both percentile and bias-corrected (BC) 95% confidence intervals. BC intervals adjust for bias in the bootstrap distribution and are recommended for products of coefficients, which typically have skewed sampling distributions [48]. A path is significant when its BC 95% CI excludes zero; this is the primary inference method for all mediation paths. We extracted individual factor scores via the regression method (mxFactorScores) for visualization of slope coupling.

##### 2.6.4.5 Coupling Model

In the coupling model, a free covariance between HVR linear slope and cognitive linear slope (hvr_s_ ↔ cog_s_) is estimated to test whether linear rates of hippocampal and cognitive decline co-vary, reported as standardized correlations:

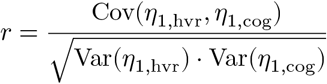

##### 2.6.4.6 Mediation Model

In the mediation model, directional paths replace the free slope covariance: the a-path (CVR → HVR slope) tests whether cardiovascular risk predicts the rate of hippocampal decline; the b-path (HVR slope → cognitive slope) captures hippocampal-cognitive coupling; the c’-path (CVR → cognitive slope) captures the direct effect bypassing hippocampal atrophy; and the indirect effect (a × b) quantifies mediation through the hippocampus. No free hvr_s_ ↔ cog_s_ covariance is estimated in the mediation model because the directional b-path already captures the slope relationship; the coupling model uses a free covariance without directional paths instead. A significant total effect is not required for valid mediation inference [41].

#### 2.6.5 Sensitivity Analyses

##### 2.6.5.1 LME Sensitivity

We conducted five sensitivity analyses within the LME framework. First, for **HC z-score specificity**, we repeated the 3-way interaction with total hippocampal volume (HC z-score, normalized against UKB norms) instead of HVR, testing whether cardiovascular risk moderation effects are specific to the hippocampus-to-ventricle ratio or generalize to hippocampal volume alone. Second, for **attrition analysis**, we compared baseline characteristics between completers (≥ 4 visits) and dropouts (< 4 visits) using t-tests (continuous) and *χ*^2^ tests (categorical), plus logistic regression predicting dropout status from baseline characteristics. Third, for **raw HVR and HC sensitivity**, we repeated the 3-way interaction using raw (un-normalized) HVR and raw HC (with intracranial volume covariate) instead of UKB z-scored values, to confirm findings are not artifacts of the normative transformation. Fourth, for **EDT moderation**, we used EDT as a moderator in a 4-way interaction (CVR × HVR × EDT × YRS). Note that EDT and YRS capture different sources of variation: EDT is a baseline moderator indexing disease stage, while YRS is the within-person time metric capturing change over follow-up; their inclusion tests whether cardiovascular risk effects on brain-cognition coupling vary by disease stage. Fifth, for **EDT Early/Middle sensitivity**, we restricted LME models to the first two EDT tertiles, excluding late-stage participants to assess stability of effects across disease stages.

##### 2.6.5.2 Amyloid-Negative CU Control

To assess whether the primary LME results are driven by AD pathology rather than cardiovascular risk, we repeated the 3-way interaction analysis in an independent sample of amyloid-negative, baseline cognitively unimpaired (A^−^/CU) participants (N = 300). Models were pooled across sex (with SEX as covariate) given the smaller sample. We also fit a linear LGCM mediation model (using years from baseline as the time metric) in this sample; results are reported in Appendix C.

### 2.7 Software

All analyses were conducted in R (R version 4.5.1 (2025-06-13)) with the following packages: lavaan [49] for MIMIC model specification, estimation (MLR + FIML), measurement invariance testing, and factor score extraction; lme4 [50] for LME growth models with bobyqa optimizer and restricted maximum likelihood (REML) estimation; OpenMx [51] for LGCM parallel process and mediation models with sequential least squares programming (SLSQP) optimizer (convergence criterion: gradient < 10^−5^); progmod [36] for EDT computation; and CVrisk [33] for FRS computation (via ADSP-PHC).

## 3 Results

### 3.1 Sample Characteristics

The LME cohort comprised 923 amyloid-positive participants (513 males, 410 females) with complete data for both FRS and CVR_mimic_ after common-sample filtering. Mean age was 73.9 ± 7.1 years (range 55– 95). The diagnostic composition included 250 cognitively unimpaired (27%), 475 MCI (51%), and 193 AD dementia (21%). Participants had a median of 4 visits (range 1–17) with mean follow-up of 4.0 years (max 17.0). Education averaged 16.1 ± 2.7 years, and 60% were APOE *ε*4 carriers. Baseline FRS was 24.2 ± 7.2 (raw); CVR_mimic_ ζ was 0.00 ± 1.00 (age-adjusted latent scores; both FRS and CVR_mimic_ z-standardized on the common sample before analysis). The LGCM cohort included 908 participants (507 males, 401 females) with ≥2 follow-ups and available EDT values. Mediation bootstrap samples comprised 507 males and 401 females (complete cases required for case-resampling). Participant inclusion and exclusion flow is shown in Figure 1; baseline demographics by sex appear in Table 1.

**Table 1:** Baseline cohort characteristics. ADNI analysis cohort compared with the UK Biobank normative reference sample. Values are mean (SD) for continuous variables, % for categorical. HC = bilateral hippocampal volume; HVR = hippocampus-to-ventricle ratio; FRS = Framingham Risk Score; APOE *ε*4+ = carriers of at least one *ε*4 allele. Z-scores derived from UK Biobank GAMLSS normative models.

| Characteristic | UKB Normative |  | ADNI Analysis Cohort |  |
| --- | --- | --- | --- | --- |
|  | Males | Females | Males | Females |
| N | 9,615 | 12,327 | 513 | 410 |
| Age, years | 64.1 (7.7) | 62.9 (7.3) | 74.9 (6.8) | 72.7 (7.3) |
| Education, years | 13.8 (2.6) | 13.8 (2.6) | 16.5 (2.7) | 15.6 (2.7) |
| HC bilateral, cm<sup>3</sup> | 6.81 (0.65) | 6.27 (0.59) | 5.50 (1.01) | 5.29 (0.91) |
| HVR (ratio) | 0.738 (0.053) | 0.763 (0.044) | 0.559 (0.119) | 0.623 (0.102) |
| HC z-score | — | — | -1.75 (1.73) | -1.43 (1.59) |
| HVR z-score | — | — | -2.27 (1.74) | -1.89 (1.38) |
| FRS, % | — | — | 21.1 (7.6) | 28.1 (4.1) |
| APOE $\epsilon 4+$, % | — | — | 60.0 | 60.9 |
| Diagnosis, $n$ (%) | — | — | | |
| CU | — | — | 116 (22.6) | 134 (32.7) |
| MCI | — | — | 281 (54.8) | 194 (47.3) |
| AD | — | — | 112 (21.8) | 81 (19.8) |

We validated UKB-derived z-scores against an amyloid-negative, cognitively unimpaired ADNI subsample within the UKB age range. HVR z-scores showed a systematic negative offset consistent with the older ADNI age distribution; HC z-scores were somewhat closer to expected values (Appendix A, Figure A1).

### 3.2 Cardiovascular Risk Characterization

#### 3.2.1 FRS Properties

In the ADNI cohort, FRS was strongly correlated with age (r = 0.326). The FRS-age partial correlation (controlling for other cardiovascular risk factors) remained substantial (r = 0.358), confirming that age dominates FRS in older adults. Variance decomposition showed age alone R^2^ = 0.107, cardiovascular risk factors alone R^2^ = 0.406, and combined R^2^ = 0.482, indicating true cardiovascular risk factors contribute modestly beyond age. A total of 48.9% of participants were at the 30% risk cap (28.1% males, 74.9% females), and 72.2% scored above 20, limiting statistical sensitivity even below the formal ceiling.

#### 3.2.2 CVR_mimic_ Validation

The MIMIC model showed good fit: CFI = 1.000, TLI = 1.020, RMSEA = 0.000, SRMR = 0.009. The structural path showed *γ* = 0.125 with R^2^ = 0.016, indicating age explained only 1.6% of cardiovascular risk variance and confirming CVR_mimic_ is largely age-free. Factor loadings appear in Table 2; visual validation in Figure 2.

**Table 2:** CVR_mimic_ model factor loadings. λ = unstandardized factor loading; Std. λ = standardized loading; SE = standard error.

| Indicator | $\lambda$ | Std. $\lambda$ | SE | p |
| --- | --- | --- | --- | --- |
| Systolic BP, z-scored | 1.000 | 0.367 | 0.000 | — |
| Hypertension, binary | 0.655 | 0.482 | 0.079 | $2.22 \times 10^{-16}$ |
| Glucose, z-scored | 0.383 | 0.141 | 0.098 | $8.77 \times 10^{-5}$ |
| Total Cholesterol, z-scored | -1.064 | -0.391 | 0.134 | $2.00 \times 10^{-15}$ |
| Creatinine, z-scored | 1.224 | 0.449 | 0.138 | $2.22 \times 10^{-16}$ |

**Figure 2:**
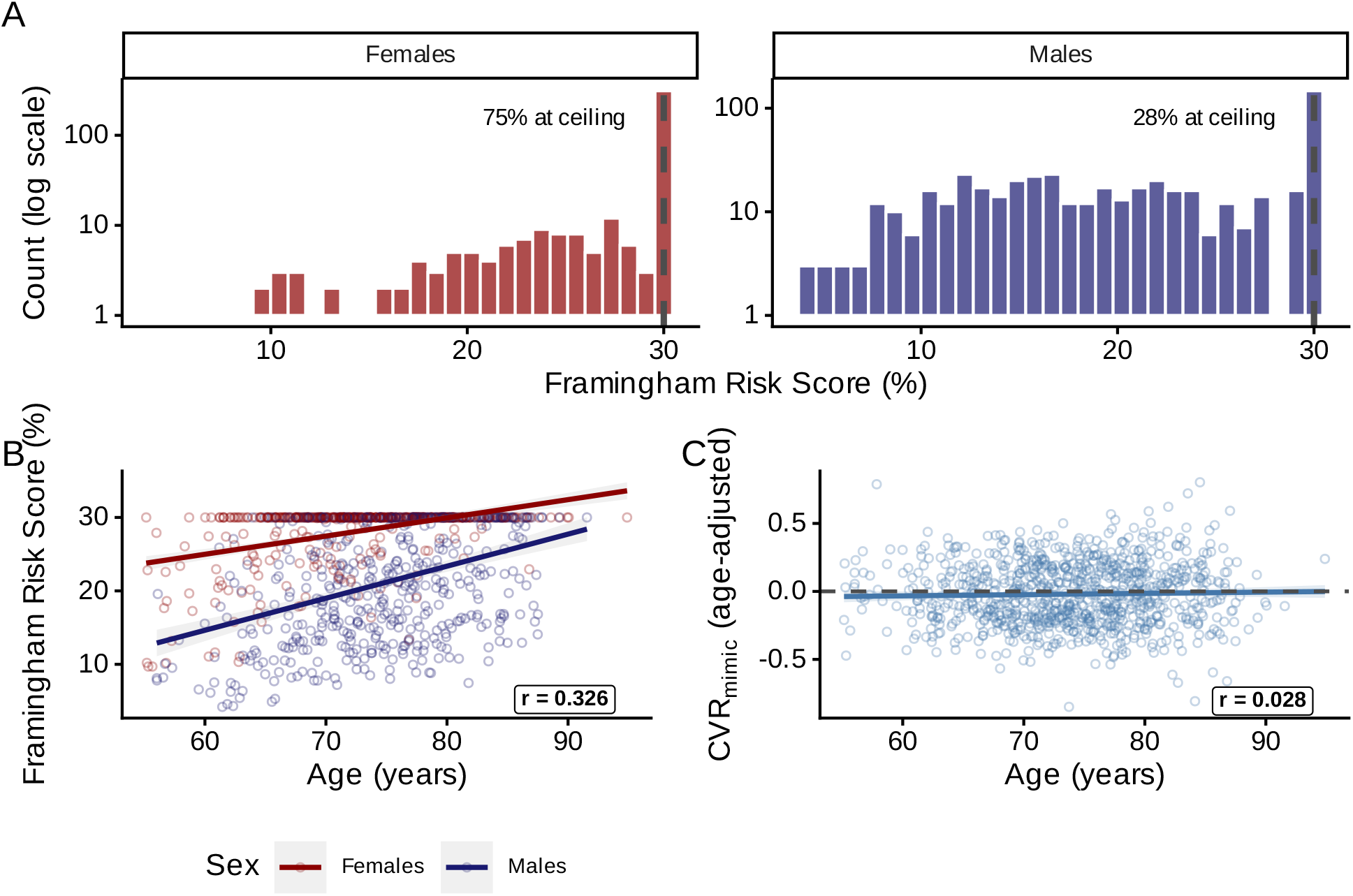
Cardiovascular risk characterization. (A) FRS distribution by sex; dashed red line marks the 30% risk ceiling. (B) FRS vs Age by sex showing strong positive correlation driven by age-weighting in the FRS formula. (C) CVR_mimic_ vs Age showing near-zero correlation, confirming successful age adjustment; dashed line marks zero.

After age-adjustment and z-standardization, CVR_mimic_ showed near-zero correlation with age (r = 0.028, by construction), moderate correlation with FRS (r = 0.12, reflecting shared CV indicators), and approximate standard normal distribution (mean = 0.00, SD = 1.00).

#### 3.2.3 Measurement Invariance

Measurement invariance of the CVR_mimic_ model was tested across sex in four steps (Appendix B, Table B1). Configural invariance was supported: CFI = 0.962, RMSEA = 0.039.

Metric (equal loadings): ΔCFI = 0.009, ΔRMSEA = −0.005 (p = 0.194); Scalar (equal intercepts): ΔCFI = 0.756, ΔRMSEA = 0.084 (p < 0.001). Scalar invariance was not achieved (|ΔCFI| > 0.01), indicating intercepts differ across sex. Metric invariance (equal loadings) was supported — sufficient for sex-stratified analyses.

EDT linearity tests and SE-constrained LGCM simulation validation are reported in Appendix B (see Appendix B, Table B2).

### 3.3 LME Results: FRS vs CVR_mimic_ 3-Way Interaction

#### 3.3.1 3-Way Interaction (FRS vs CVR_mimic_)

Full covariate effects (Age, Education, APOE *ε*4, cardiovascular risk main effect, HVR z-score) are reported in Appendix B (Tables B3 and B4).

##### Key pattern

FRS produced significant 3-way interactions (YRS × CVR × HVR) in males (Memory, Language, Executive Function) and females (Memory, Language, Executive Function) — all six models, all surviving FDR correction. CVR_mimic_ showed only one of six significant interactions (none of the six surviving FDR), consistent with FRS effects being primarily driven by age-weighting rather than true cardiovascular risk (see Figure 3).

**Figure 3:**
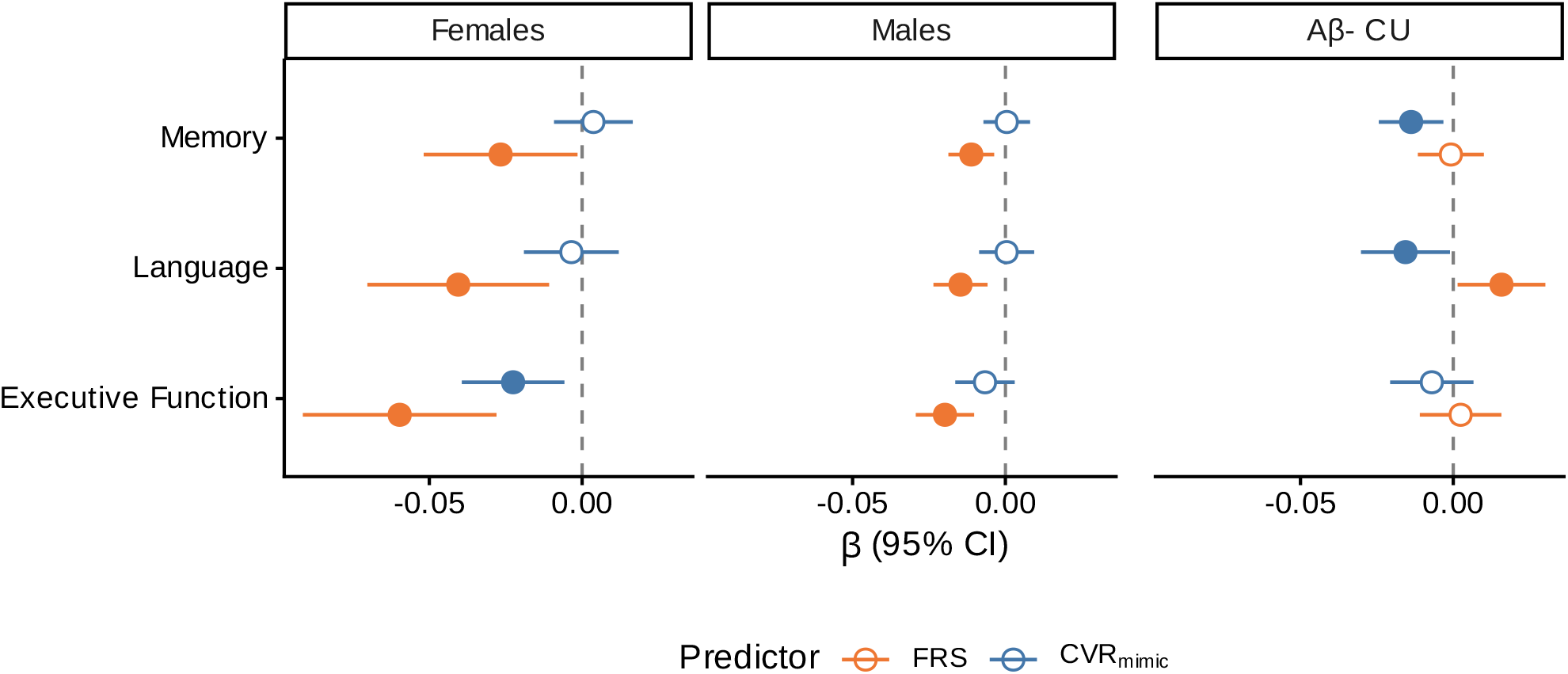
LME 3-way interaction forest plot. FRS vs CVR_mimic_ effects (YRS × CVR × HVR z-score → Cognition) by sex and cognitive domain, with an amyloid-negative cognitively unimpaired (A^−^/CU) control panel (pooled sex). Filled points indicate p < 0.05; white-filled points indicate p ≥ 0.05. Error bars show 95% CI.

##### Males

FRS × HVR × YRS was significant for Memory (*β* = −0.0112, p = 0.004 [*p*_FDR_ = 0.005]), Language (*β* = −0.0147, p = 0.001 [*p*_FDR_ = 0.002]), and Executive Function (*β* = −0.0198, p < 0.001 [*p*_FDR_ < 0.001]). CVR_mimic_ showed no significant interactions (all *p* ≥ 0.179).

##### Females

FRS effects were also significant across all three domains, with larger magnitudes: Memory (*β* = −0.0267, p = 0.038 [*p*_FDR_ = 0.038]), Language (*β* = −0.0405, p = 0.008 [*p*_FDR_ = 0.009]), and Executive Function (*β* = −0.0597, p < 0.001 [*p*_FDR_ < 0.001]). CVR_mimic_ reached significance only for Executive Function (*β* = −0.0226, p = 0.008 [*p*_FDR_ = 0.051, n.s.]).

FRS *β* values were on average 4.6-fold larger in absolute magnitude than CVR_mimic_ *β* values, consistent with age-weighting amplifying apparent FRS effects. The largest significant FRS 3-way effect (Female Executive Function, *β* = −0.0597) implies that over 5 years, a 1-SD increase in FRS is associated with a −0.2986-unit cumulative divergence in the HVR-cognition association.

FDR correction did not change FRS conclusions. For CVR_mimic_, 0 of 1 uncorrected effects survived FDR.

#### 3.3.2 Formal Coefficient Comparison (LME)

To formally compare FRS and CVR_mimic_ 3-way interaction coefficients, we computed clustered bootstrap Δ*β* with bias-corrected 95% CIs.

##### Males

Δ*β* values were negative across all three domains, reaching significance for Language (Δ*β* = −0.0151, 95% CI [−0.0306, −0.0018]) but not Memory (Δ*β* = −0.0116, 95% CI [−0.0254, 0.0007]) or Executive Function (Δ*β* = −0.0131, 95% CI [−0.0284, 0.0002]).

##### Females

Δ*β* values were likewise negative. Memory reached significance (Δ*β* = −0.0304, 95% CI [−0.0700, −0.0010]), whereas Language (Δ*β* = −0.0370, 95% CI [−0.0882, 0.0097]) and Executive Function (Δ*β* = −0.0371, 95% CI [−0.0861, 0.0138]) did not.

The A^−^/CU control analysis reversed this pattern: CVR_mimic_ reached FDR significance while FRS did not (Figure 3, panel C; full results in Appendix C).

### 3.4 LGCM Results

#### 3.4.1 Growth Parameters

LGCM models used EDT as the time metric with individually-varying time scores, fit separately per sex and cognitive domain.

Model fit appears in Table 3; standard fit indices (CFI, TLI, RMSEA) are not computable with individually-varying time scores, so adequacy was assessed via −2LL, AIC, BIC, and nested LRTs.

**Table 3:** LGCM mediation model fit indices. Sex-stratified. −2LL = −2 log-likelihood; AIC = Akaike information criterion; BIC = Bayesian information criterion.

| Mediation Model Fit |  |  |  |  |
| --- | --- | --- | --- | --- |
|  | Females |  | Males |  |
| | FRS | $\text{CVR}_{\text{mimic}}$ | FRS | $\text{CVR}_{\text{mimic}}$ |
| **Memory** |  |  |  |  |
| -2LL | 30,153.2 | 30,400.6 | 35,257.3 | 35,176.8 |
| AIC | 30,225.2 | 30,472.6 | 35,329.3 | 35,248.8 |
| BIC | 30,369.0 | 30,616.3 | 35,481.5 | 35,401.1 |
| **Language** |  |  |  |  |
| -2LL | 30,480.5 | 30,728.8 | 36,252.0 | 36,165.7 |
| AIC | 30,552.5 | 30,800.8 | 36,324.0 | 36,237.7 |
| BIC | 30,696.3 | 30,944.6 | 36,476.3 | 36,390.0 |
| **Executive Function** |  |  |  |  |
| -2LL | 30,930.9 | 31,174.0 | 36,735.1 | 36,645.7 |
| AIC | 31,002.9 | 31,246.0 | 36,807.1 | 36,717.7 |
| BIC | 31,146.6 | 31,389.8 | 36,959.3 | 36,869.9 |

#### 3.4.2 Slope Coupling

LGCM parallel process models tested whether HVR and cognitive linear decline rates co-vary (latent slope coupling, hvr_s_ ↔ cog_s_) along the EDT disease timeline.

In males, linear slope correlations (*r*) ranged from 0.285 (Memory) to 0.824 (Executive Function; max p = 0.011). In females, linear slope correlations (*r*) ranged from 0.149 (Memory) to 0.396 (Executive Function; max p = 0.202). In males, faster hippocampal decline was significantly associated with steeper cognitive decline across all three domains; in females, the direction was consistent but only Executive Function reached significance, likely reflecting lower statistical power in the smaller female subsample. This coupling supports the b-path assumption in the mediation framework: HVR decline predicts cognitive decline. Latent factor scores from the parallel process model confirm this pattern (see Figure 4).

**Figure 4:**
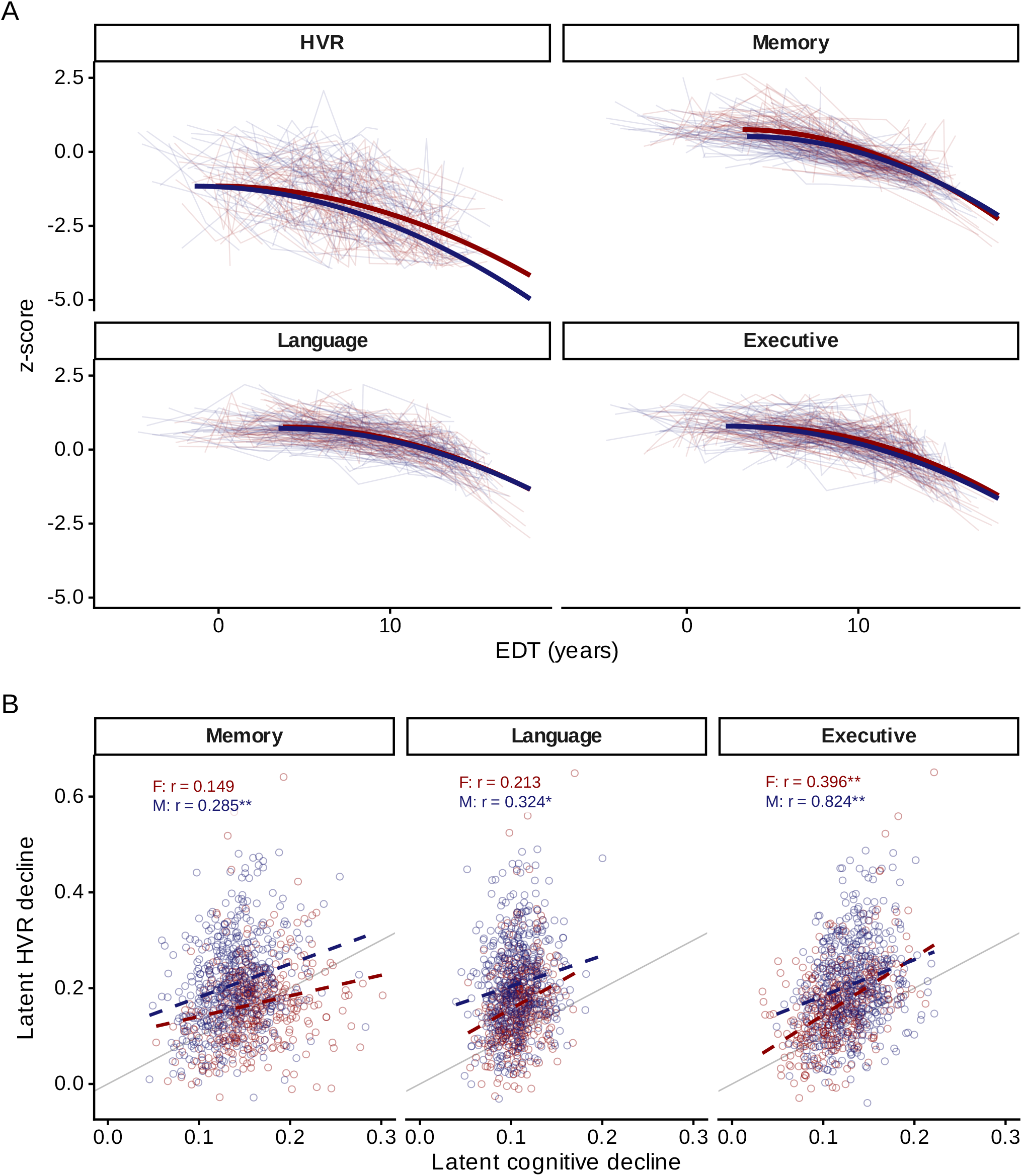
HVR–cognitive latent slope coupling. Females in red, Males in blue throughout. A. Individual trajectories (random 20% subsample) with population mean curves (bold; including fixed quadratic curvature where selected by LRT). 2×2 facets: HVR, Memory, Language, Executive. B. Latent slope coupling: latent cognitive decline rate (x) vs latent HVR decline rate (y) from factor scores (positive = faster decline); dashed lines show linear trends per sex; grey line shows 1:1 reference. Correlation estimates from the SEM (* p < .05, ** p < .01, *** p < .001).

#### 3.4.3 Mediation: FRS vs CVR_mimic_

LGCM mediation models tested whether cardiovascular risk effects on cognitive decline operate through hippocampal atrophy, comparing FRS and CVR_mimic_ as predictors (see Figure 5).

**Figure 5:**
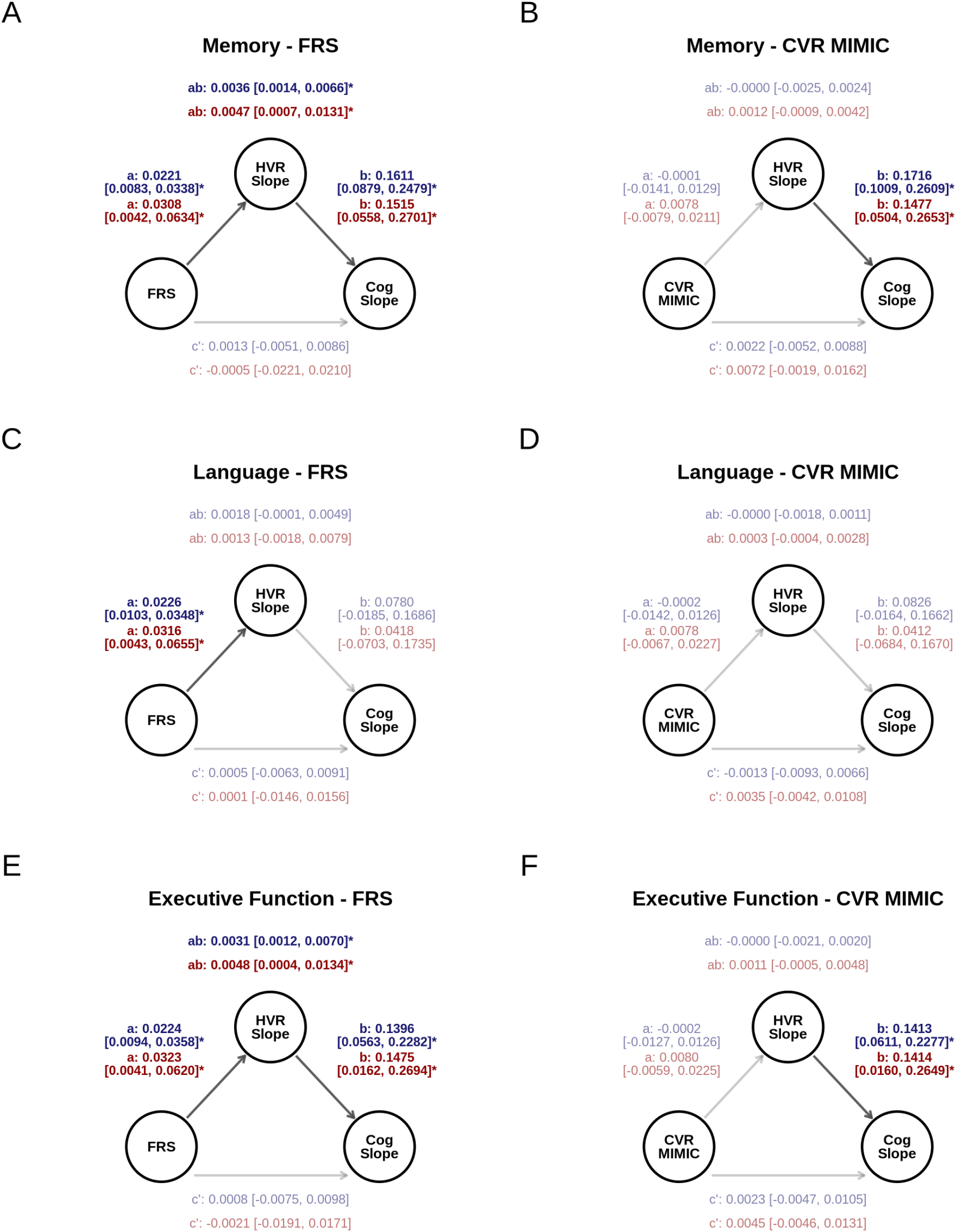
LGCM mediation path diagrams. Rows: Memory, Language, Executive Function. Columns: FRS, CVR_mimic_. Each panel shows Male (blue) and Female (red) path estimates. Bold = significant (boot-strap 95% CI excludes zero); faded = non-significant. Star (*) and bold indicate CI excludes zero. 95% bootstrap CIs shown for all paths.

Mediation path diagrams showing a-, b-, c’-path estimates and bootstrap CIs appear in Figure 5.

##### FRS vs CVR_mimic_ across all 12 models

The a-path (CVR → HVR slope) showed a clear predictor dissociation: FRS significantly predicted hippocampal decline in all six models (bootstrap 95% CIs excluding zero), whereas CVR_mimic_ did not in any — indicating that FRS “effects” on hippocampal decline are largely consistent with age-weighting. By contrast, the b-path (HVR slope → cognitive slope) was significant for four of six FRS and four of six CVR_mimic_ models, confirming that hippocampal-cognitive coupling persists regardless of predictor. The FRS indirect effect (a × b) was significant in four of six models; CVR_mimic_ indirect effects in none of the six.

##### Males

FRS predicted hippocampal atrophy across all three domains (a-path *β* range 0.0221 to 0.0226, all bootstrap 95% CIs excluding zero); CVR_mimic_ did not (all |*β*| < 0.01). Hippocampal decline in turn predicted cognitive decline (b-path *β* range 0.078 to 0.161, two of three CIs excluding zero), and the FRS indirect effect was significant for two of three domains (proportion mediated 73%–80%). No CVR_mimic_ indirect effects reached significance.

##### Females

The same a-path dissociation held: FRS was significant across all three domains (*β* range 0.0308 to 0.0323), CVR_mimic_ was not (all |*β*| < 0.01). B-path estimates were comparable to males (*β* range 0.042 to 0.151, two of three CIs excluding zero), and FRS indirect effects reached significance in two of three domains (proportion mediated 90%–full mediation); CVR_mimic_ indirect effects reached significance in none of the three. No CVR_mimic_ direct effects (c’) reached significance.

Complete path estimates, bootstrap CIs, and direct (c’) effects appear in Figure 5.

#### 3.4.4 Formal a-Path Comparison (LGCM)

Paired bootstrap with shared-seed resampling was used to formally compare FRS and CVR_mimic_ a-path coefficients (individual estimates shown in Figure 5 above).

The paired difference Δ_a_ = a_FRS_ − a_CVR_ quantifies how much larger the FRS a-path is than the CVR_mimic_ a-path; a significant positive Δ_a_ confirms FRS is the stronger predictor of hippocampal atrophy rate.

##### Males

Δ_a_ was significantly positive across all three domains — Memory (Δ_a_ = 0.02221, 95% CI [0.00737, 0.03627]), Language (Δ_a_ = 0.02283, 95% CI [0.00914, 0.03759]), and Executive Function (Δ_a_ = 0.02258, 95% CI [0.00912, 0.03753]) — confirming a consistent FRS advantage.

##### Females

Δ_a_ was positive across all three domains (Memory Δ_a_ = 0.02300, 95% CI [−0.00357, 0.05044]; Language Δ_a_ = 0.02374, 95% CI [−0.00271, 0.05420]; Executive Function Δ_a_ = 0.02425, 95% CI [−0.00085, 0.05413]), but all three 95% BC CIs narrowly included zero.

Paired bootstrap confirmed the a-path dissociation in three of six sex × domain models: Δ_a_ (FRS − CVR_mimic_) was positive in every case, with 95% BC CIs excluding zero across all three male domains; no female domain reached significance. Male Δ_a_ estimates were consistent across domains (mean Δ_a_ = 0.0225), indicating a stable FRS advantage regardless of cognitive outcome. Female Δ_a_ estimates were approximately 5% larger (mean Δ_a_ = 0.0237), suggesting the FRS-CVR_mimic_ dissociation is more pronounced in females. These formal tests upgrade the qualitative dissociation (FRS a-path significant in all six models, CVR_mimic_ in none) to a quantitative difference: FRS is a significantly stronger predictor of hippocampal atrophy rate than CVR_mimic_ within the mediation framework.

Sensitivity analyses — including HC z-score comparisons, attrition analysis, raw (un-normalized) concordance checks, EDT moderation, SE-constrained LGCM validation, and A^−^/CU control analysis — are reported in Appendix C.

## 4 Discussion

### 4.1 Summary of Findings

The apparent impact of cardiovascular risk on hippocampal-cognitive trajectories depends on whether the risk measure retains its age component. The LGCM a-path dissociated: FRS predicted hippocampal decline in all six models (Figure 5), CVR_mimic_ in none, indicating age-weighting drives the FRS-hippocampal link. The b-path (hippocampal-cognitive coupling) remained significant for both FRS (four of six) and CVR_mimic_ (four of six), so hippocampal atrophy predicts cognitive decline regardless of CVR measurement [52–54]. The LME 3-way interaction (FRS × HVR × YRS) was significant across all three male and all three female domains (all surviving FDR), while CVR_mimic_ reached significance only in one of three female domains (Executive Function only) and none of the three male, suggesting residual cardiovascular burden may modulate HVR-cognition independently of accelerated atrophy.

### 4.2 FRS Age-Confounding and CVR_mimic_

CVR_mimic_ successfully removes the age component from cardiovascular risk (near-zero age correlation; Figure 2 C), supporting its use as an age-independent index. Its moderate FRS correlation reflects shared cardiovascular indicators while confirming distinct constructs: one age-confounded, one age-orthogonal.

The FRS formula heavily weights age [5], and in older samples age dominates the score [6,7]. FRS values in ADNI are compressed into the upper range (Figure 2, panels A-B), limiting score variance and statistical sensitivity, particularly in females. When age is partialled out (CVR_mimic_), the indirect pathway disappears: the a-path becomes null, suggesting the FRS-hippocampal association is primarily age-driven. This does not invalidate FRS clinically; age is itself a potent marker of cumulative vascular exposure [9]. However, FRS was originally validated for adults aged 30–74 [5], and ADNI participants substantially exceed this range and often have prevalent cardiovascular disease, raising questions about FRS validity in this population.

The negative total cholesterol loading (Table 2) likely reflects widespread statin use in this elderly clinical sample: treated individuals have lower cholesterol despite elevated underlying risk, inverting the expected loading direction. Prior FRS-brain reports in elderly samples [1–4] therefore likely conflate age-driven and cardiovascular effects that are difficult to disentangle without age-adjustment.

### 4.3 Dissociating Moderation from Mediation

The two modelling frameworks test complementary questions. The LME 3-way interaction (YRS × CVR × HVR → Cognition) tests moderation: does cardiovascular risk modify HVR-cognition over time? The LGCM a-path (CVR → HVR slope; Figure 5) tests mediation: does cardiovascular risk affect cognition through hippocampal decline?

For CVR_mimic_, the LGCM a-path was null across all six models, suggesting age-adjusted cardiovascular risk does not accelerate hippocampal atrophy. Yet the LME 3-way interaction was significant in one of three female domains (Executive Function) only, with none of the three in males. This pattern represents moderation without mediation: CVR_mimic_ does not drive structural decline (a-path null) but may modify how hippocampal integrity relates to cognitive performance over time, consistent with a cognitive reserve framework [55–57]. The A^−^/CU analysis strengthens this: with AD pathology absent, CVR_mimic_ reached FDR significance while FRS did not (Figure 3), suggesting a cardiovascular reserve effect masked by amyloid-driven hippocampal decline in the A^+^ sample.

The HVR-cognition coupling (b-path) was consistent across both cardiovascular risk predictors, in line with prior LGCM coupling studies [41,42].

### 4.4 Sex Differences

CVR_mimic_ LME effects reached significance in none of the three male domains and one of three female domains (Executive Function). The female CVR_mimic_ Executive finding is the only CVR_mimic_ LME effect in either sex, consistent with vascular burden selectively impairing executive function in MCI [58]; replication in independent samples is needed. Female FRS variance compression limits statistical sensitivity, so FRS findings in females may be primarily age-driven. Sex differences in cardiovascular risk profiles [22,23], differ-ential APOE *ε*4 effects, and hormonal influences on vascular regulation may all contribute to this asymmetry. Covariate effects (age, education, APOE *ε*4) appear in Appendix B (Appendix B, Tables B3 and B4).

The sex asymmetry extended to the age covariate. Despite GAMLSS age-normalization of HVR z-scores, age → HVR intercept and slope paths remained significant in all six male mediation models (intercept −0.049 to −0.065, all *p* < 6.5e-07; slope all *p* < 2.1e-06), indicating that UKB-trained normative models did not fully absorb age effects in older ADNI males (consistent with the transfer offset in Appendix A). Female age effects on HVR were uniformly non-significant (all *p* > 0.18). Male age coefficients were larger in FRS models (mean |*β*| = 0.065) than CVR_mimic_ models (0.050), consistent with FRS and age competing for shared variance. Baseline age also predicted the cognitive intercept in all twelve models (all *p* < 9.2e-03), justifying its retention as a covariate.

### 4.5 HVR-Cognitive Coupling

The HVR slope → Cognitive slope relationship (b-path) was consistent regardless of whether FRS or CVR_mimic_ was the predictor (Figure 5). The b-path was significant in four of six FRS and four of six CVR_mimic_ models, supporting HVR decline as a reliable marker of cognitive decline [52,53] and extending evidence that HVR captures hippocampal integrity more sensitively than raw volume [26,31,59].

Parallel process coupling showed a complementary pattern: significant in all three male models but only Executive Function in females (one of three). This sex asymmetry likely reflects lower power in the smaller female subsample, since mediation b-paths (which condition on a predictor) were significant in four of six models for both sexes. Language mediation b-paths were non-significant for both sexes; parallel process coupling reached Language significance in males but not females, consistent with later-declining domains showing less variance at this disease stage. The hippocampal-cognitive coupling is not an artifact of cardiovascular risk measurement; it persists across predictor specifications and analysis frameworks.

### 4.6 Methodological Considerations

The fixed-quadratic LGCM captures population-level curvature but constrains individual quadratic variation, so slope estimates primarily reflect average rates of change. This makes the a-path null finding conservative: any individual-level non-linear CVR_mimic_ effects on hippocampal trajectories would require a fully random quadratic model to detect, further strengthening the FRS age-weighting interpretation.

EDT linearity tests confirmed non-linear trajectories (Appendix B, Table B2), motivating the fixed-quadratic specification [60]. Fully random quadratic models failed to converge due to near-zero variance. EDT accommodates heterogeneous disease progression but is sensitive to the disease model’s assumptions [36].

Sensitivity analyses (Appendix C) showed HC z-score effects were weaker than HVR effects (Appendix C, Table C1), attrition patterns suggest observed effects may underestimate true relationships (Appendix C, Table C4), and raw-volume results were concordant with z-scored analyses.

### 4.7 Clinical Implications

In clinical trials targeting cardiovascular risk reduction for dementia prevention, FRS remains a valid summary of overall vascular burden, including the age-driven component that reflects cumulative exposure [5,9]. When the question concerns cardiovascular effects independent of aging, age-adjusted measures such as CVR_mimic_ provide a necessary complement [8,21]. HVR is a more sensitive neuroimaging endpoint than hippocampal volume alone, and its consistent cognitive coupling supports use as a surrogate outcome in intervention studies. The isolated CVR_mimic_ Executive Function finding (one of six tests, non-significant after FDR) requires replication before clinical interpretation.

### 4.8 Limitations

This study used a single cohort (ADNI), and replication in other aging samples is needed. UKB-trained GAMLSS normative models showed a systematic negative z-score offset in the ADNI comparable subsample due to age distribution mismatch; within-ADNI relative comparisons remain valid, but absolute z-scores should be interpreted cautiously across cohorts. CVR_mimic_ indicators were limited to available cardiovascular risk factor data and measured cross-sectionally; richer or longitudinal indicators could refine the construct. FRS values in this elderly cohort cluster near the 30% 10-year ceiling, particularly in females, so null CVR_mimic_ findings may partly reflect insufficient variance after age removal rather than a true absence of cardiovascular effects. The A^−^/CU control analysis showed a reversed FRS/CVR_mimic_ pattern, suggesting generalizability beyond the AD continuum, but the smaller pooled-sex subsample limits this inference.

ADAS-13 and MMSE items contribute to both EDT and the Memory/Language composites; this overlap is mitigated by (1) the bifactor CFA extracting domain-specific variance, diluting any single shared item’s contribution, and (2) EDT entering as a time metric rather than an outcome. Executive Function has minimal item overlap (Appendix B, Figure B1).

Coefficient-comparison tests share the resampling framework of mediation inference, without independent validation. Individually-varying time scores preclude CFI/TLI/RMSEA; model adequacy was assessed via information criteria and nested LRTs. The MIMIC model used 5 indicators; richer models may yield different CVR_mimic_ values. Bootstrap mediation CIs (N = 507 males, 401 females) may be liberal with these modest sample sizes.

### 4.9 Conclusions

Whether cardiovascular risk appears to drive hippocampal-cognitive decline depends on how it is measured. FRS produces significant indirect effects via hippocampal decline; age-adjusted CVR_mimic_ does not. Because age both marks cumulative vascular exposure and drives neurodegeneration [9], FRS-hippocampal associations primarily reflect age. The A^−^/CU analysis (Figure 3; Appendix C) shows the converse: with amyloid burden removed, CVR_mimic_ reached FDR significance while FRS did not, consistent with a reserve-like mechanism. Hippocampal-cognitive coupling persists across both measures, supporting HVR as a reliable biomarker target.

Future directions include replication in independent aging cohorts, longitudinal cardiovascular measurement, and intervention trials. The MIMIC age-adjustment framework also applies to composite scores where a constituent independently predicts the outcome.

## Data Availability

Data were obtained from two access-controlled repositories. (1) The Alzheimer's Disease Neuroimaging Initiative (ADNI; https://adni.loni.usc.edu), accessed via the LONI Image and Data Archive following registration and approval of a Data Use Agreement. (2) The UK Biobank (https://www.ukbiobank.ac.uk), accessed under Number 45551; UK Biobank data were used to derive the normative GAMLSS z-score models applied in this study.

https://www.ukbiobank.ac.uk

## Ethics Approval Statement

Data used in preparation of this article were obtained from the ADNI database. ADNI was approved by the Institutional Review Boards of all participating institutions. Written informed consent was obtained from all participants.

## Author Contributions

**S.F-L**.: Conceptualization, Methodology, Software, Formal analysis, Investigation, Data curation, Visualization, Writing – original draft. **S.V**.: Conceptualization, Writing – review & editing. **D.L.C**.: Supervision, Project administration, Writing – review & editing. All authors read and approved the final manuscript.

## Acknowledgments

Data used in preparation of this article were obtained from the Alzheimer’s Disease Neuroimaging Initiative (ADNI) database (adni.loni.usc.edu). As such, the investigators within the ADNI contributed to the design and implementation of ADNI and/or provided data but did not participate in analysis or writing of this report. A complete listing of ADNI investigators can be found at: <http://adni.loni.usc.edu/wp-content/uploads/how_to_apply/ADNI_Acknowledgement_List.pdf>

The authors used Claude (Anthropic) for language editing of the manuscript text. The tool was not used for data collection, statistical analysis, scientific interpretation, or generation of figures. The authors are solely responsible for the design, analysis, and conclusions of this work.

## Data Availability Statement

This research was conducted using ADNI data (adni.loni.usc.edu). Data are available to qualified researchers through the ADNI data sharing platform. Analysis code is available at https://github.com/soffiafdz/hvr-cvr-mediation.

