## Appendices for "Cardiovascular risk and hippocampal-cognitive coupling in Alzheimer’s disease"

#### A Appendix A: Normative Transfer Validation

For transfer validation, we identified a comparable ADNI subsample of 293 amyloid-negative cognitively unimpaired participants (57% female) within the UKB age range. Despite matching on amyloid status and cognitive normality, the ADNI comparable subsample was approximately 8 years older on average (mean 71.3 vs 63.4 years; Figure A1 A), concentrating in the upper tail of the UKB age distribution. HVR z-scores showed a systematic negative offset (mean = -0.96, SD = 1.14, 95% CI [-1.09, -0.83]), while HC z-scores were closer to expected values (mean = -0.41, SD = 1.16; Figure A1 B). Age-bin-specific calibration (Figure A1, panel B) showed mean z-scores near zero in younger bins with increasingly negative offset in older bins, consistent with the age mismatch.

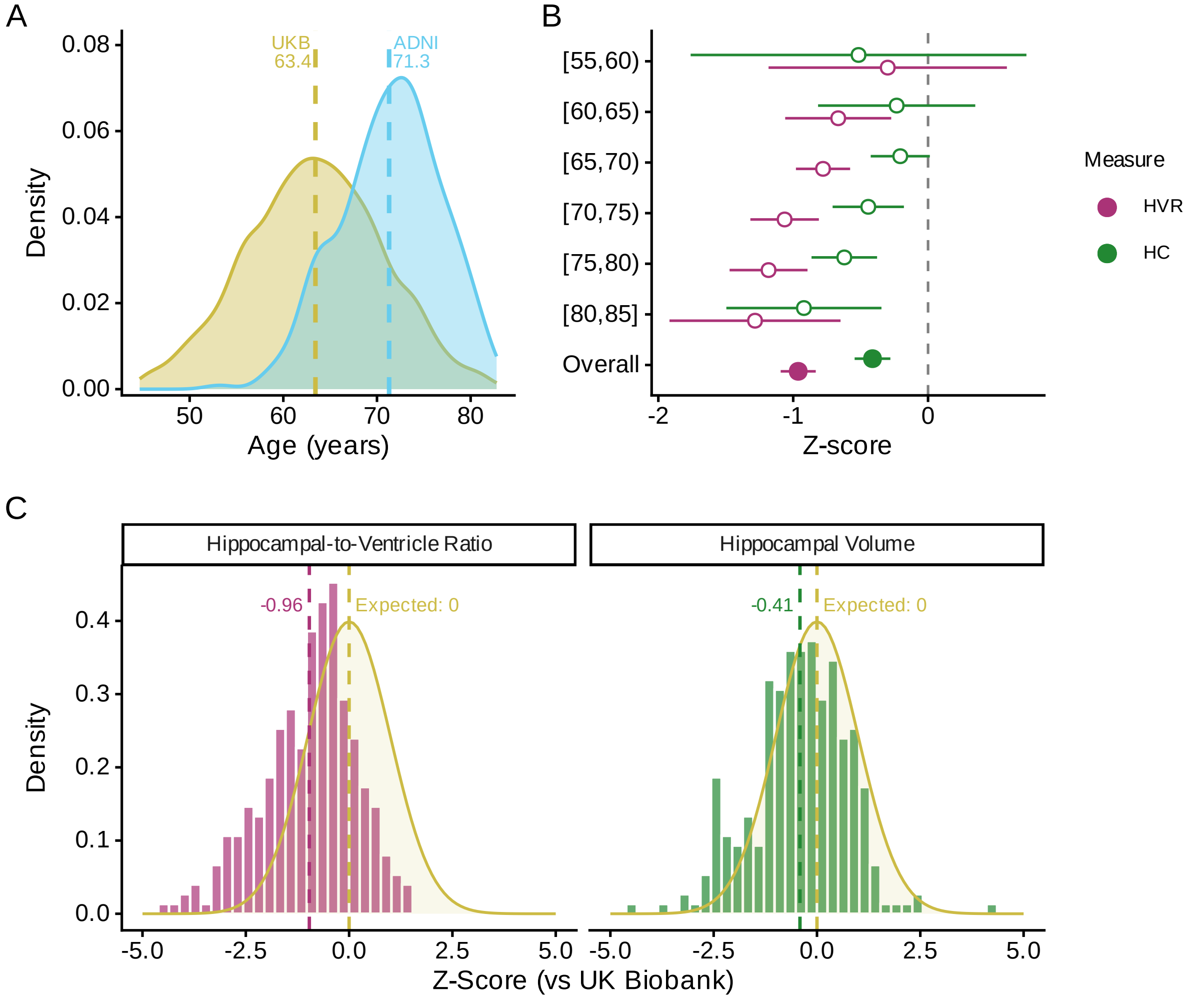


Figure A1: **Normative transfer validation.** (A) Age distributions: UK Biobank (UKB, yellow) vs ADNI comparable subsample (cyan); dashed lines indicate sample means. (B) Forest plot of HVR (purple) and HC (green) z-scores (mean ± 95% CI) by age bin; reference line at z = 0 indicates perfect calibration; open circles = age bins, filled circles = overall. (C) Z-score distributions for HVR (left) and HC (right) compared to the expected N(0,1) (yellow).

### B Appendix B: Methodological Validations

#### B.1 $\text{CVR}_{\text{mimic}}$ Measurement Invariance

Measurement invariance of the $\text{CVR}_{\text{mimic}}$ model was tested across sex in four steps (configural, metric, scalar, strict; Table B1).

Table B1: **$\text{CVR}_{\text{mimic}}$ measurement invariance.** Configural = same structure; Metric = equal loadings; Scalar = equal intercepts; Strict = equal residuals. $\Delta$CFI $< 0.01$ and $\Delta$RMSEA $< 0.015$ indicate acceptable invariance.

Measurement Invariance Across Sex

| | Model Fit | | | | | | Comparison | | |
|---|---|---|---|---|---|---|---|---|---|
| Level | $\chi^2$ | df | CFI | TLI | RMSEA | SRMR | $\Delta$CFI | $\Delta$RMSEA | Decision |
| Configural | 14.094 | 6 | 0.962 | 0.875 | 0.039 | 0.021 | — | — | — |
| Metric | 20.673 | 10 | 0.954 | 0.907 | 0.034 | 0.025 | 0.009 | -0.005 | HOLD |
| Scalar | 222.891 | 14 | 0.198 | -0.146 | 0.118 | 0.072 | 0.756 | 0.084 | REJECT |
| Strict | 196.316 | 19 | 0.000 | -0.176 | 0.119 | 0.102 | — | — | — |

#### B.2 EDT Linearity and Simulation Validation

The linear time specification was validated via LME likelihood ratio tests comparing linear vs quadratic EDT across all sex-domain models (see Table B2).

Table B2: **EDT linearity validation.** LME models testing linear vs quadratic EDT specification. LRT tests whether adding a quadratic EDT term significantly improves fit. $\Delta$AIC = AIC(linear) − AIC(quadratic); positive values favor quadratic. EDT = Estimated Disease Time.

EDT Linearity Validation

| | | | Likelihood Ratio Test | | |
|---|---|---|---|---|---|
| | N | $\Delta$AIC | $\chi^2$ | df | p |
| Memory | 4,872 | 2,470.79 | 2,472.79 | 1 | < 2.2e-16 |
| Language | 4,872 | 1,469.40 | 1,471.40 | 1 | 6.42e-322 |
| Executive Function | 4,868 | 1,343.35 | 1,345.35 | 1 | 1.58e-294 |

All three models showed significant quadratic EDT terms (Memory, Language, Executive Function; max $\chi^2$ = 2472.8). Large test statistics reflect substantial power afforded by sample size and repeated observations. A fixed-quadratic specification was adopted: quadratic variance fixed to near zero with freely estimated quadratic means, capturing population-level curvature without individual-level quadratic variation.

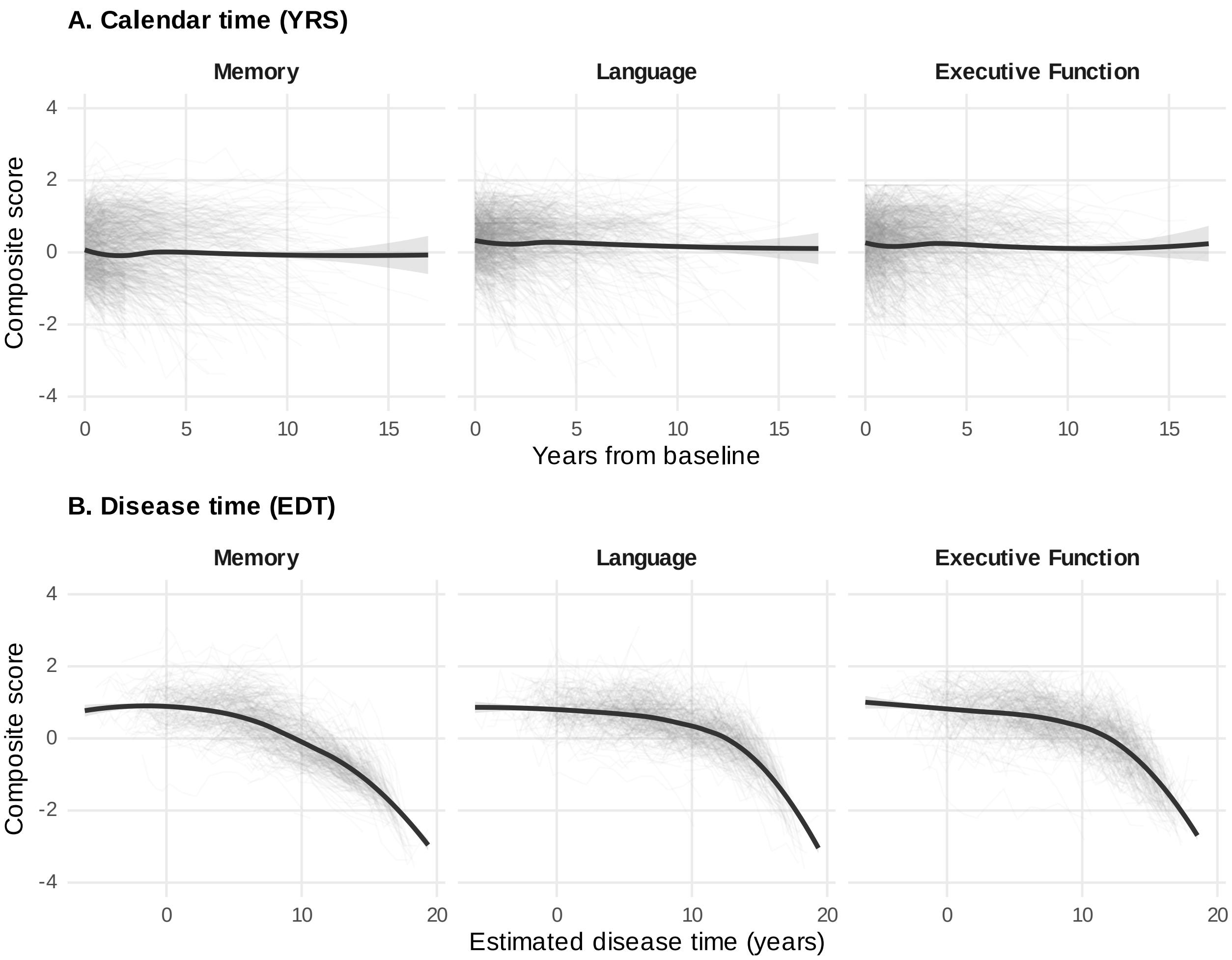


Figure B1: **Cognitive trajectories under calendar time versus estimated disease time.** Composite scores (Memory, Language, Executive Function) plotted against years from baseline (A) and estimated disease time (B). Under calendar time, the population-average trajectory appears flat because participants at different disease stages are pooled at each follow-up; under EDT, disease-stage alignment reveals the expected monotonic cognitive decline. Lines show loess smooths with 95% CI bands; individual trajectories in light gray. Y-axes are shared across panels for direct comparison; x-axes differ because the two metrics have different natural scales.

#### B.3 Standard Error-Constrained LGCM Validation

A Monte Carlo simulation (500 replications per condition at N = 200 and N = 500) compared SE-constrained LGCM (residual variance fixed to $SE^2$) against free estimation. Parameters were calibrated from the observed ADNI LGCM results (see Methods). To assess robustness, the HVR effect and slope variance were varied at 0.5×, 1×, and 1.5× their observed values, yielding 9 conditions.

At the base condition (observed ADNI parameters), the SE-constrained approach converged in 500 of 500 replications at N = 200 (free: 500 of 500) with bias = -5.9e-06 (free: 3.2e-05), RMSE = 0.0092 (free: 0.0093), and 95% CI coverage = 93.2% (free: 93.2%). At N = 500, bias was 7.8e-05 (free: 1.1e-04), RMSE = 0.0062 (free: 0.0062), and coverage = 93.8% (free: 93.4%).

Across all 9 grid conditions, maximum absolute bias was 5.9e-04, coverage ranged from 93.2% to 96.2% for SE-constrained and 93.2% to 96.6% for free estimation. Both approaches showed comparable performance across all realistic parameter combinations, validating the SE-constrained specification used in the primary analyses. The equivalence between methods under ADNI-calibrated conditions confirms that fixing residual variance to $SE^2$ neither introduces bias nor distorts inference, while incorporating known measurement information from the ADSP-PHC scoring model.

#### B.4 LME Covariate Effects

Full covariate effects from the sex-stratified LME 3-way interaction models are presented for the FRS model (Table B3) and the $CVR_{mimic}$ model (Table B4).

Table B3: **FRS LME covariate effects.** Sex-stratified. Bold indicates $p < 0.05$.

Covariate Effects (FRS Model)

| | Females | | | Males | | |
|---|---|---|---|---|---|---|
| | $\beta$ | SE | p | $\beta$ | SE | p |
| **Memory** | | | | | | |
| Education | **0.0685** | **0.0123** | **$5.22 \times 10^{-8}$** | **0.0510** | **0.0093** | **$7.33 \times 10^{-8}$** |
| APOE e4+ | **-0.3029** | **0.0697** | **$1.80 \times 10^{-5}$** | **-0.2252** | **0.0513** | **$1.40 \times 10^{-5}$** |
| FRS | **0.2233** | **0.1022** | **0.029** | -0.0088 | 0.0391 | 0.823 |
| HVR z-score | **0.1894** | **0.0278** | **$1.92 \times 10^{-11}$** | **0.1229** | **0.0136** | **$2.22 \times 10^{-16}$** |
| **Language** | | | | | | |
| Education | **0.0639** | **0.0103** | **$1.67 \times 10^{-9}$** | **0.0482** | **0.0088** | **$6.38 \times 10^{-8}$** |
| APOE e4+ | -0.0430 | 0.0587 | 0.464 | -0.0450 | 0.0484 | 0.353 |
| FRS | 0.0893 | 0.0918 | 0.331 | 0.0157 | 0.0384 | 0.684 |
| HVR z-score | **0.1419** | **0.0265** | **$1.15 \times 10^{-7}$** | **0.1282** | **0.0137** | **$2.22 \times 10^{-16}$** |
| **Executive Function** | | | | | | |
| Education | **0.0790** | **0.0123** | **$4.39 \times 10^{-10}$** | **0.0461** | **0.0106** | **$1.74 \times 10^{-5}$** |
| APOE e4+ | -0.1152 | 0.0700 | 0.100 | -0.1070 | 0.0587 | 0.069 |
| FRS | 0.0717 | 0.1084 | 0.509 | -0.0719 | 0.0458 | 0.117 |
| HVR z-score | **0.1919** | **0.0310** | **$1.07 \times 10^{-9}$** | **0.1077** | **0.0162** | **$6.62 \times 10^{-11}$** |

Table B4: **$CVR_{mimic}$ LME covariate effects.** Sex-stratified. Bold indicates $p < 0.05$.

Covariate Effects ($CVR_{mimic}$ Model)

| | Females | | | Males | | |
|---|---|---|---|---|---|---|
| | $\beta$ | SE | p | $\beta$ | SE | p |
| **Memory** | | | | | | |
| Education | **0.0645** | **0.0125** | **$4.47 \times 10^{-7}$** | **0.0494** | **0.0094** | **$1.95 \times 10^{-7}$** |
| APOE e4+ | **-0.2860** | **0.0702** | **$5.72 \times 10^{-5}$** | **-0.2381** | **0.0513** | **$4.55 \times 10^{-6}$** |
| $CVR_{mimic}$ | -0.0410 | 0.0538 | 0.446 | -0.0410 | 0.0402 | 0.308 |
| HVR z-score | **0.1854** | **0.0207** | **$2.22 \times 10^{-16}$** | **0.1290** | **0.0131** | **$2.22 \times 10^{-16}$** |
| **Language** | | | | | | |
| Education | **0.0596** | **0.0106** | **$3.11 \times 10^{-8}$** | **0.0464** | **0.0087** | **$1.48 \times 10^{-7}$** |
| APOE e4+ | -0.0288 | 0.0591 | 0.627 | -0.0606 | 0.0480 | 0.207 |
| $CVR_{mimic}$ | 0.0097 | 0.0493 | 0.844 | -0.0721 | 0.0387 | 0.063 |
| HVR z-score | **0.1353** | **0.0198** | **$1.93 \times 10^{-11}$** | **0.1280** | **0.0132** | **$2.22 \times 10^{-16}$** |
| **Executive Function** | | | | | | |
| Education | **0.0728** | **0.0125** | **$1.27 \times 10^{-8}$** | **0.0428** | **0.0106** | **$6.18 \times 10^{-5}$** |
| APOE e4+ | -0.1059 | 0.0702 | 0.132 | **-0.1244** | **0.0583** | **0.033** |
| $CVR_{mimic}$ | -0.0032 | 0.0580 | 0.955 | **-0.1225** | **0.0464** | **0.009** |
| HVR z-score | **0.1879** | **0.0232** | **$2.81 \times 10^{-15}$** | **0.1161** | **0.0156** | **$3.24 \times 10^{-13}$** |

### C Appendix C: Sensitivity Analyses

All sensitivity checks converge on the same pattern: FRS effects trace to age-weighting, $CVR_{mimic}$ effects are null or isolated, and the HVR–cognition coupling (b-path) is consistent. Main findings are stable across alternative brain measures (HC volume), raw scores, attrition adjustments, EDT specification checks, and an amyloid-negative CU control analysis. Full details follow.

#### C.1 LME Sensitivity

##### C.1.1 HC z-score (Total Hippocampal Volume)

HC z-score effects were generally weaker than HVR z-score effects, with fewer significant 3-way interactions, supporting the hippocampus-to-ventricle ratio as a more sensitive brain aging biomarker. The LME 3-way interaction was repeated with HC z-score instead of HVR; results appear in Table C1, with covariate effects in Table C2 and Table C3.

Table C1: **HC z-score sensitivity analysis.** LME 3-way interaction effects (Years × cardiovascular risk × HC z-score) for total hippocampal volume. FRS and $CVR_{mimic}$ compared side-by-side. Bold indicates $p < 0.05$.

| | Females | | | | Males | | | |
|---|---|---|---|---|---|---|---|---|
| | $\beta$ | SE | p | $p_{FDR}$ | $\beta$ | SE | p | $p_{FDR}$ |
| **Memory** | | | | | | | | |
| FRS | 0.0044 | 0.0109 | 0.686 | 0.686 | -0.0072 | 0.0039 | 0.070 | 0.139 |
| $CVR_{mimic}$ | 0.0050 | 0.0055 | 0.365 | 0.603 | 0.0034 | 0.0040 | 0.402 | 0.603 |
| **Language** | | | | | | | | |
| FRS | -0.0104 | 0.0129 | 0.422 | 0.506 | -0.0041 | 0.0047 | 0.383 | 0.506 |
| $CVR_{mimic}$ | 0.0009 | 0.0066 | 0.888 | 0.888 | 0.0059 | 0.0048 | 0.215 | 0.603 |
| **Executive Function** | | | | | | | | |
| FRS | **-0.0349** | **0.0139** | **0.012** | 0.070 | **-0.0115** | **0.0051** | **0.023** | 0.070 |
| $CVR_{mimic}$ | **-0.0150** | **0.0072** | **0.037** | 0.222 | -0.0015 | 0.0051 | 0.777 | 0.888 |

The FRS × HC × Years interaction was significant for Male Executive Function and Female Executive Function, whereas $CVR_{mimic}$ × HC × Years reached significance only for Female Executive Function. HC z-score showed weaker effects than HVR z-score, supporting use of the ratio measure.

Significant 3-way interactions emerged in three of twelve models for HC z-score versus nine of twelve for HVR z-score. The weaker HC effects support the specificity of HVR: capturing ventricular expansion alongside hippocampal atrophy provides greater sensitivity to cardiovascular risk moderation.

Table C2: **FRS HC z-score covariate effects.** Sex-stratified. Bold indicates $p < 0.05$.

HC Covariate Effects (FRS Model)

| | Females | | | Males | | |
|---|---|---|---|---|---|---|
| | $\beta$ | SE | p | $\beta$ | SE | p |
| **Memory** | | | | | | |
| Education | **0.0702** | **0.0120** | **$1.11 \times 10^{-8}$** | **0.0525** | **0.0092** | **$2.28 \times 10^{-8}$** |
| APOE e4+ | **-0.2161** | **0.0686** | **0.002** | **-0.1844** | **0.0512** | **$3.48 \times 10^{-4}$** |
| FRS | **0.2144** | **0.0774** | **0.006** | 0.0501 | 0.0339 | 0.140 |
| **Language** | | | | | | |
| Education | **0.0649** | **0.0105** | **$1.78 \times 10^{-9}$** | **0.0504** | **0.0090** | **$3.94 \times 10^{-8}$** |
| APOE e4+ | 0.0117 | 0.0603 | 0.846 | -0.0240 | 0.0502 | 0.634 |
| FRS | **0.1680** | **0.0695** | **0.016** | 0.0547 | 0.0338 | 0.106 |
| **Executive Function** | | | | | | |
| Education | **0.0816** | **0.0129** | **$7.27 \times 10^{-10}$** | **0.0479** | **0.0111** | **$2.00 \times 10^{-5}$** |
| APOE e4+ | -0.0415 | 0.0740 | 0.575 | -0.1069 | 0.0617 | 0.084 |
| FRS | **0.2246** | **0.0845** | **0.008** | 0.0145 | 0.0411 | 0.724 |

Table C3: **$CVR_{mimic}$ HC z-score covariate effects.** Sex-stratified. Bold indicates $p < 0.05$.

HC Covariate Effects ($CVR_{mimic}$ Model)

| | Females | | | Males | | |
|---|---|---|---|---|---|---|
| | $\beta$ | SE | p | $\beta$ | SE | p |
| **Memory** | | | | | | |
| Education | **0.0665** | **0.0122** | **$8.97 \times 10^{-8}$** | **0.0507** | **0.0092** | **$6.21 \times 10^{-8}$** |
| APOE e4+ | **-0.2044** | **0.0695** | **0.003** | **-0.1994** | **0.0510** | **$1.07 \times 10^{-4}$** |
| $CVR_{mimic}$ | -0.0483 | 0.0427 | 0.258 | -0.0577 | 0.0350 | 0.100 |
| **Language** | | | | | | |
| Education | **0.0604** | **0.0107** | **$2.77 \times 10^{-8}$** | **0.0486** | **0.0090** | **$9.13 \times 10^{-8}$** |
| APOE e4+ | 0.0103 | 0.0610 | 0.866 | -0.0390 | 0.0498 | 0.434 |
| $CVR_{mimic}$ | 0.0079 | 0.0384 | 0.836 | **-0.1098** | **0.0347** | **0.002** |
| **Executive Function** | | | | | | |
| Education | **0.0747** | **0.0129** | **$1.61 \times 10^{-8}$** | **0.0450** | **0.0110** | **$4.96 \times 10^{-5}$** |
| APOE e4+ | -0.0575 | 0.0741 | 0.438 | **-0.1250** | **0.0609** | **0.041** |
| $CVR_{mimic}$ | -0.0010 | 0.0463 | 0.983 | **-0.1258** | **0.0420** | **0.003** |

##### C.1.2 Attrition Analysis

Baseline characteristics were compared between completers (≥4 visits) and dropouts (<4 visits); results appear in Table C4.

Table C4: **Attrition analysis.** Baseline characteristics of completers (≥4 visits) vs dropouts (<4 visits). Bold rows indicate $p < 0.05$. Completers = participants with sufficient visits for quadratic trend estimation.

| | Completers | | Dropouts | | | | |
|---|---|---|---|---|---|---|---|
| | Mean/% | SD | Mean/% | SD | Δ | $\chi^2$/t | p |
| Sex (% Male) | 56.17 | — | 54.30 | — | 1.88 | 0.21 | $6.44 \times 10^{-1}$ |
| Age | 73.74 | 6.95 | 74.25 | 7.43 | -0.51 | -0.99 | $3.20 \times 10^{-1}$ |
| Education | 16.06 | 2.76 | 16.23 | 2.68 | -0.17 | -0.91 | $3.65 \times 10^{-1}$ |
| FRS | **-0.04** | **1.02** | **0.22** | **0.89** | **-0.26** | **-3.94** | $\mathbf{8.92 \times 10^{-5}}$ |
| HVR (z-score) | -2.03 | 1.54 | -2.26 | 1.71 | 0.23 | 1.95 | $5.14 \times 10^{-2}$ |
| HC (z-score) | -1.54 | 1.67 | -1.76 | 1.68 | 0.22 | 1.87 | $6.16 \times 10^{-2}$ |
| APOE $\varepsilon$4 (%) | **58.07** | — | **65.52** | — | **-7.45** | **4.30** | $\mathbf{3.80 \times 10^{-2}}$ |

Significant baseline differences between completers and dropouts were observed in FRS, APOE $\varepsilon$4 carrier status (all $p < 0.05$; see Table C4).

Logistic regression predicting dropout identified 2 significant predictors — FRS (OR = 1.52, 95% CI [1.26, 1.84]); APOE $\varepsilon$4 carrier status (OR = 1.37, 95% CI [1.02, 1.86]). Dropouts had higher baseline FRS (M = 0.22) than completers (M = -0.04), consistent with selective loss of higher-risk participants — this may attenuate observed cardiovascular risk effects.

##### C.1.3 Raw Brain Measure Sensitivity

The LME 3-way interaction was repeated using raw (un-normalized) HVR and raw HC (with total intracranial volume [TIV] covariate), with FRS as predictor, full-sample, no sex stratification, and no Years$^2$ term.

Raw HVR was significant for all three domains: Memory ($\beta$ = -0.0162, p = 0.003), Language ($\beta$ = -0.0133, p = 0.042), Executive Function ($\beta$ = -0.0200, p = 0.005). Raw HC (+TIV) reached significance only for Executive Function ($\beta$ = -0.0169, p = 0.017). Raw HVR × FRS was significant for Memory, Language, Executive Function, confirming normative z-scoring does not drive primary findings.

##### C.1.4 EDT Moderation of LME Effects

A 4-way interaction (cardiovascular risk × HVR × EDT × Years) tested whether cardiovascular risk effects vary by disease stage.

FRS × HVR × EDT (Male): significant for Memory ($\beta$ = -0.0005, p = 0.008); Language ($\beta$ = -0.0009,

$p < 0.001$); Executive Function ($\beta$ = -0.0008, $p < 0.001$). FRS × HVR × EDT (Female): significant for Language ($\beta$ = -0.0023, $p < 0.001$); Executive Function ($\beta$ = -0.0023, p = 0.002). $CVR_{mimic}$ × HVR × EDT (Male): not significant (min p = 0.299). $CVR_{mimic}$ × HVR × EDT (Female): significant for Language ($\beta$ = -0.0050, p = 0.005). Limited EDT moderation suggests cardiovascular risk effects on HVR-cognition coupling are relatively stable across disease stages.

##### C.1.5 EDT Stage Sensitivity

Analyses were restricted to early and middle EDT tertiles (excluding late-stage) to confirm effect stability.

HVR z-score: Memory ($\beta$ = 0.0038, p = 0.750, 185.6% change); Language ($\beta$ = -0.0044, p = 0.687, 80.7% change); Executive Function ($\beta$ = -0.0008, p = 0.944, 97.0% change) (1/3 concordant with full-sample). HC z-score: Memory ($\beta$ = 0.0271, p = 0.018, 7.4% change); Language ($\beta$ = 0.0088, p = 0.392, 56.7% change); Executive Function ($\beta$ = 0.0140, p = 0.212, 38.8% change) (3/3 concordant with full-sample).

#### C.2 Amyloid-Negative CU Control

##### C.2.1 LME 3-Way Interaction

The LME 3-way interaction was repeated in 300 $A^-$/CU participants (pooled sex, SEX as covariate; see Methods). Results appear in Table C5.

Table C5: **$A^-$/CU LME 3-way interaction.** FRS vs $CVR_{mimic}$ effects (YRS × CVR × HVR z-score → Cognition) in amyloid-negative, cognitively unimpaired participants. Pooled sex with SEX as covariate. Bold indicates $p < 0.05$. FDR correction applied within each predictor (3 tests).

| | FRS | | | | $CVR_{mimic}$ | | | |
|---|---|---|---|---|---|---|---|---|
| | $\beta$ | SE | p | $p_{FDR}$ | $\beta$ | SE | p | $p_{FDR}$ |
| Executive Function | 0.0024 | 0.0068 | 0.723 | 0.888 | -0.0070 | 0.0070 | 0.313 | 0.313 |
| Language | **0.0158** | **0.0073** | **0.032** | **0.095** | **-0.0156** | **0.0074** | **0.036** | **0.054** |
| Memory | -0.0008 | 0.0055 | 0.888 | 0.888 | **-0.0138** | **0.0054** | **0.011** | **0.032** |

FRS did not survive FDR correction in any domain (none of the three; Memory (p = 0.888); Language (p = 0.032); Executive Function (p = 0.723)), whereas $CVR_{mimic}$ reached significance in one of three domains after FDR correction (Memory ($\beta$ = -0.0138, p = 0.011 [$p_{FDR}$ = 0.032]); Language ($\beta$ = -0.0156, p = 0.036 [$p_{FDR}$ = 0.054, n.s.]); Executive Function ($\beta$ = -0.0070, p = 0.313)). This complementary reversal — FRS significant in $A^+$ but not after FDR in $A^-$/CU; $CVR_{mimic}$ not significant after FDR in $A^+$ but significant in $A^-$/CU — supports the interpretation that FRS effects are carried by the age component of the score, which co-varies with AD pathology (see main text, Figure 2).

##### C.2.2 LGCM Mediation

The $A^{-}$/CU LGCM used a linear growth model with YRS time metric, pooled sexes, free residuals, and Wald/Sobel inference (see Methods; N = 300, analytic N = 260).

Table C6: **$A^{-}$/CU LGCM mediation paths.** Linear LGCM mediation in amyloid-negative, cognitively unimpaired participants. a-path = predictor → HVR slope; b-path = HVR slope → cognitive slope; indirect = a × b. Bold indicates $p < 0.05$. Inference via Wald/Sobel tests (no bootstrap).

| | FRS | | $CVR_{mimic}$ | |
|---|---|---|---|---|
| | $\beta$ | p | $\beta$ | p |
| **Executive Function** | | | | |
| a (cardiovascular risk →HVR slope) | 0.0048 | 0.582 | 0.0020 | 0.804 |
| b (HVR →Cog slope) | **0.2834** | **0.036** | **0.2826** | **0.034** |
| Indirect (a ×b) | 0.0014 | 0.597 | 0.0006 | 0.806 |
| **Language** | | | | |
| a (cardiovascular risk →HVR slope) | 0.0053 | 0.546 | 0.0021 | 0.792 |
| b (HVR →Cog slope) | 0.3072 | 0.052 | 0.3034 | 0.051 |
| Indirect (a ×b) | 0.0016 | 0.571 | 0.0006 | 0.794 |
| **Memory** | | | | |
| a (cardiovascular risk →HVR slope) | 0.0051 | 0.560 | 0.0024 | 0.759 |
| b (HVR →Cog slope) | **0.3561** | **0.003** | **0.3619** | **0.003** |
| Indirect (a ×b) | 0.0018 | 0.571 | 0.0009 | 0.761 |

Neither FRS nor $CVR_{mimic}$ predicted HVR slope in any of the 6 models (all a-path p = 0.546; Table C6). This contrasts sharply with the primary $A^{+}$ analysis, where FRS predicted hippocampal decline in all six models (see main text, Figure 4): removing amyloid pathology eliminates the FRS a-path, confirming that FRS-hippocampal associations require AD-related neurodegeneration rather than reflecting direct cardiovascular effects on the hippocampus. The a-path estimates themselves are near zero (range 0.002–0.005), indicating this is a genuinely null effect rather than a power limitation.

The b-path (HVR slope → cognitive slope) was significant for Memory ($\beta \approx 0.359$, p = 0.003 to p = 0.003) and Executive Function ($\beta \approx 0.283$, p = 0.034 to p = 0.036), and borderline for Language (p = 0.051 to p = 0.052), confirming that hippocampal-cognitive coupling persists in the absence of amyloid burden. This invariance across both $A^{+}$ and $A^{-}$/CU samples and across both cardiovascular risk predictors supports HVR decline as a general marker of neurodegeneration-cognition coupling, independent of AD pathology or cardiovascular risk measurement.

No indirect effects reached significance (all Sobel $p > 0.5$), consistent with null a-paths. The overall pattern — null a-paths with preserved b-paths for both FRS and $CVR_{mimic}$ — closes the age-confounding argument: FRS mediates through hippocampal decline only when age co-varies with AD-driven atrophy ($A^{+}$), not in the general aging population ($A^{-}$/CU).
